# Huntington’s Disease Integrated Staging System (HD-ISS): A Novel Evidence-Based Classification System For Staging

**DOI:** 10.1101/2021.09.01.21262503

**Authors:** Sarah J. Tabrizi, Scott Schobel, Emily C. Gantman, Alexandra Mansbach, Beth Borowsky, Pavlina Konstantinova, Tiago A. Mestre, Jennifer Panagoulias, Christopher A. Ross, Maurice Zauderer, Ariana P. Mullin, Klaus Romero, Sudhir Sivakumaran, Emily C. Turner, Jeffrey D. Long, Cristina Sampaio, on behalf of the Huntington’s Disease Regulatory Science Consortium (HD-RSC)

**Author notes:** **Corresponding authors:** Professor Sarah J Tabrizi, MD, FRCP, PhD, FMedSci., Director, Huntington’s Disease Centre, and Joint-Head, Department of Neurodegenerative Disease, UCL Queen Square Institute of Neurology. Principal Investigator, UK Dementia Research Institute at University College London., Professor Cristina Sampaio MD, PhD, Chief Clinical Officer, CHDI Management/CHDI Foundation Inc, and Clinical Pharmacology Laboratory, Faculdade de Medicina de Lisboa, Portugal. joint first authors (Sarah J. Tabrizi, Scott Schobel).

## Abstract

**Background:** Despite the monogenic autosomal dominant nature of Huntington’s disease (HD), the current research paradigm is still based on overt clinical phenotypes and does not address disease pathobiology and biomarkers that are evident decades before functional decline. A new research framework is needed to standardize clinical research and enable interventional studies earlier in the course of HD.

**Methods:** The HD Regulatory Science Consortium (HD-RSC), a precompetitive Critical Path Institute initiative that includes 37 member organizations, created the Regulatory Science Forum working group (RSF), which includes industry and academic representatives. To generate a new evidenced-based HD Integrated Staging System (HD-ISS) using a formal consensus methodology, the RSF considered prognostic biomarkers, signs, and symptoms of HD, and performed empirical data analysis. We used observational data to calculate healthy-control-based landmark variable cut-offs for Stage classification and to internally validate the framework.

**Findings:** The HD-ISS starts with Stage 0, which comprises individuals with ≥ 40 cytosine-adenine-guanine repeats (CAG) in the huntingtin gene (*HTT*), before detectable indications of disease. We concluded that detectable HD progression is verified with measurable indicators of underlying pathophysiology (Stage 1), proceeds to a detectable clinical phenotype (Stage 2), and continues to a decline in function (Stage 3). Operationally, individuals can be unambiguously classified into Stages 1-3 based on CAG-independent thresholds of landmark assessments. Both cross-sectional status and longitudinal HD-ISS Stage progress align with HD natural history, and Stage transitions accelerate as CAG increase.

**Interpretation:** The HD-ISS encompasses the full course of HD starting at birth, defined by the presence of the genetic expansion. This new framework aims to standardize language for clinical research and its immediate use will enable further validation. The HD-ISS provides structure to harmonize clinical study populations and facilitates the clinical assessment of interventions earlier in HD to prevent or slow disease progression.

**Funding:** CHDI Foundation Inc.

## INTRODUCTION

Huntington’s disease (HD; OMIM #143100) is a genetic, autosomal dominant, neurodegenerative disease.^1^ While there is biological certainty that individuals who inherit a pathogenic expansion of CAG in *HTT* will eventually develop the signs and symptoms of HD, clinical practice still revolves around diagnosis based on clinical criteria that emerge well after the disease process is known to be underway.^2,3^ In the last decade, the scientific community has become acutely aware of the need to change the research paradigm to recognize the earlier phases of the disease spanning the period between birth and clinical motor diagnosis.^4-7^

The concept of clinical diagnosis predates the discovery of *HTT*; it was created to identify HD cases based on clinical certainty before it was possible to test for the presence of the expansion of *HTT* CAG, and, importantly, before our knowledge of HD-related pathobiological changes that develop many decades before observable clinical signs.^8,9^ The Diagnostic Confidence Level (DCL) item, based on motor features included in the 1999 Unified Huntington’s Disease Rating Scale (UHDRS™), was an attempt to standardize the clinical motor diagnosis concept and became widely used in research.^10,11^ DCL has been used with Shoulson and Fahn staging as the default process to classify HD participants in clinical studies.^12^

However, like the diagnosis of Alzheimer’s disease (AD) at the onset of dementia, clinical diagnosis of HD at DCL = 4 (DCL4) is a later-stage event rather than indicative of the beginning of the biological disease process. The entirety of the disease course before DCL4 has been designated by various terms, “pre-symptomatic,” “premanifest,” or “prodromal,” which have taken on different meanings, leading to non-standardized case definitions in observational studies and clinical trials.^13-18^ For example, in the last two decades there have been foundational observational studies that contribute decisively to what is currently known about HD before clinical motor diagnosis – TRACK-HD, Track On-HD, PREDICT-HD, IMAGE-HD, Registry, Enroll-HD, and more recently HD-YAS.^13-21^ However, these studies each established distinct methods, including differently-adjusted products of age and CAG,^4^ to select and categorize participant disease progression. The lack of standardization makes it difficult to describe and compare populations across studies. Recently, a task force of the Movement Disorder Society proposed a standardization of the terminology to refine clinical diagnostic criteria; however, that initiative was grounded solely in clinical criteria, excluding genetic and disease progression biomarkers for its structure.^22^

The progress of clinical research and development in the early, pre-DCL4 phases of HD requires an integrated system of classification and staging that incorporates all available data, including biomarkers, which will allow the conduct of clinical trials earlier in the disease course, data sharing with uniform case definitions, and validation of outcome measures for homogenous populations. In particular, such a system will facilitate the development of clinical trials aimed at slowing disease progression, which may need to be initiated early in the biological course of the disease for maximal benefit.

The Huntington’s Disease Regulatory Science Consortium (HD-RSC^23^) is dedicated to building regulatory science strategies and creating new tools and methods to accelerate and de-risk the development and approval of HD therapeutics (see supplementary material). The HD-RSC comprises 37 members from the biopharmaceutical industry, academia, and nonprofit and patient-advocacy organizations. The HD-RSC created the Regulatory Science Forum (RSF) as a working group to develop strategies to enable HD clinical trials before clinical motor diagnosis.

To address the issues above, the HD-RSC tasked our group, the RSF, to develop a framework that defines HD using biological criteria and to establish an evidence-based staging system, the HD-ISS. The RSF voting body is composed of ten consortium representatives from industry and academia. We received input on the HD-ISS from regulatory agency representatives and the HD patient community, and the full HD-RSC formally endorsed the HD-ISS.

## METHODS

### Formal Consensus Methodology

We, the RSF, adopted a modified version of the NIH conference consensus method^24^ in which decisions were informed by rigorous evidence gathering, and empirical data was used to develop statistical models and validate the resulting staging system. We served as the expert panel and required ≥80% favorable votes for majority adoption of proposals. The conclusions and their evidentiary basis were formally reviewed by the HD-RSC Coordinating Committee. In addition, informal feedback was incorporated from regulatory agency partners and from the HD Coalition for Patient Engagement (HD-COPE), an international patient advocacy collaboration.

### Participants in the Analysis

We used four publicly available data sets: Enroll-HD (4th periodic data set),^18^ IMAGE-HD,^25,26^ PREDICT-HD,^14,27-29^ and TRACK-HD^7,13,21^ (Table S5, S7, <35 CAG: *N* = 4636 with 13,300 longitudinal study visits; 40-50 CAG (excluding juvenile HD): *N* = 1107 with 3176 longitudinal visits with imaging). A larger dataset drawn from the four studies of *N* = 12,152 participants with 36-50 CAG without imaging was used for the reduced penetrance analysis.

### Framework Development

#### Biological HD Definition and Penetrance

Development of a biological HD definition based on molecular genetics required demonstrating the size of the *HTT* CAG expansion that is deterministic, i.e., fully penetrant. We considered the rates of penetrance reported in the literature and carried out new statistical analyses to assess the probability of developing symptomatic disease in an expected life span. To estimate the population-average probability of DCL4 over time, we fit longitudinal logistic regression models by CAG.

### HD Staging System

#### Conceptual model for stepwise progression of HD

Clinical stages classify clusters of patients into broad groups based on shared prognostic features, expected outcomes, and required treatments.^30^ HD progression models informed the sequence of events from HD inception (birth) to death to identify critical disease-stage transitions, which we operationalized by defining landmark assessments and respective cut-off values.

#### Landmark Selection

The literature review generated an exhaustive list of candidate biomarkers and assessments (collectively, “assessments”) used in HD clinical research. We only considered assessments evaluated in studies with longitudinal design and sample size >100, and prognostic of disease progression (Tables S2 and S3). We selected two landmarks each for Stages 1-3, applying an either/or rule,^31^ and gave preference to assessments representing broader constructs and less affected by language and practice effects.

#### Cut-offs Based on Controls

To define cut-off values for the chosen landmarks, we conducted new data analysis (see supplementary material). Cut-offs for each landmark variable (Stages 1-3) are the quantiles associated with the 5% tail area of the variable’s distribution in control data over time, which were estimated using quantile regression for repeated measures (Figure S1).

### Internal validation of the Staging system

Participants with CAG ≥ 40 were scored for eligibility for each stage by determining whether their observed scores exceeded the cut-offs for the assessments (0 = ineligible, 1 = eligible). Eligibility for a stage was met if either or both cut-offs were exceeded. Stage assignment (classification) was determined by summing the number of eligibilities. The proportion of monotone patterns dictated by the staging system was tallied. Kendall’s tau-a was used to index the extent to which an individual progressed through the stages over time (the ordinal correlation between the individual’s stages and their visits).^32^ The continuation-ratio model for repeated measures^33^ was used to estimate the probability of being in a stage over time as a function of CAG. Missing data was handled with multiple imputation via a method that also allowed for bootstrapped confidence intervals (CIs).^34^ Additional details of the statistical methods can be found in the supplementary material.

## RESULTS

In accordance with the consensus methodology, we voted on the answers to each question below. All votes were unanimous.

### Biological HD Definition and Penetrance

#### What size *HTT* CAG expansion is necessary and sufficient for HD pathogenesis?

While the *HTT* glutamine tract is polymorphic in the general population, the HD genetic continuum comprises individuals with an expanded CAG tract (≥27 repeats) that is pathological and/or carries an increased propensity to transmit an enlarged CAG expansion to their offspring (Table 1).^35-38^ We concluded from published studies of family pedigrees and our analysis below (Figure 1) that ≥40 CAG in exon 1 of *HTT* on chromosome 4 has a positive predictive value for clinical manifestations of HD superior to 94%, and therefore can be considered causal.^39,40^

**Table 1.**
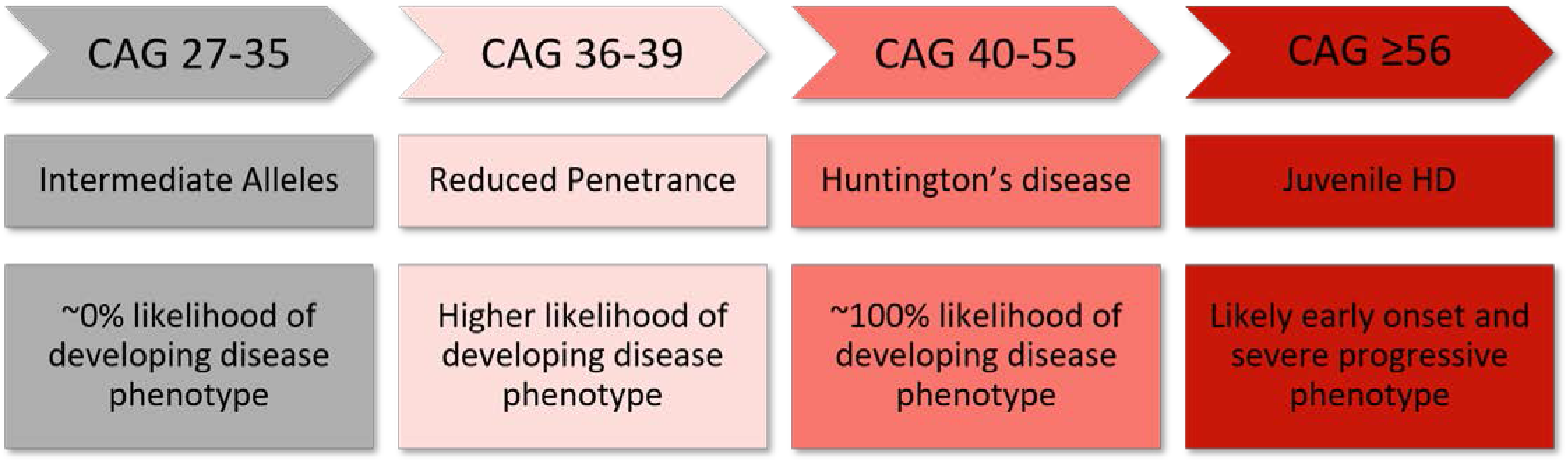
HD genetic continuum. Individuals with CAG > 26 have an increased propensity to transmit an enlarged CAG expansion; the likelihood of developing the disease phenotype increases with CAG.

**Figure 1.**
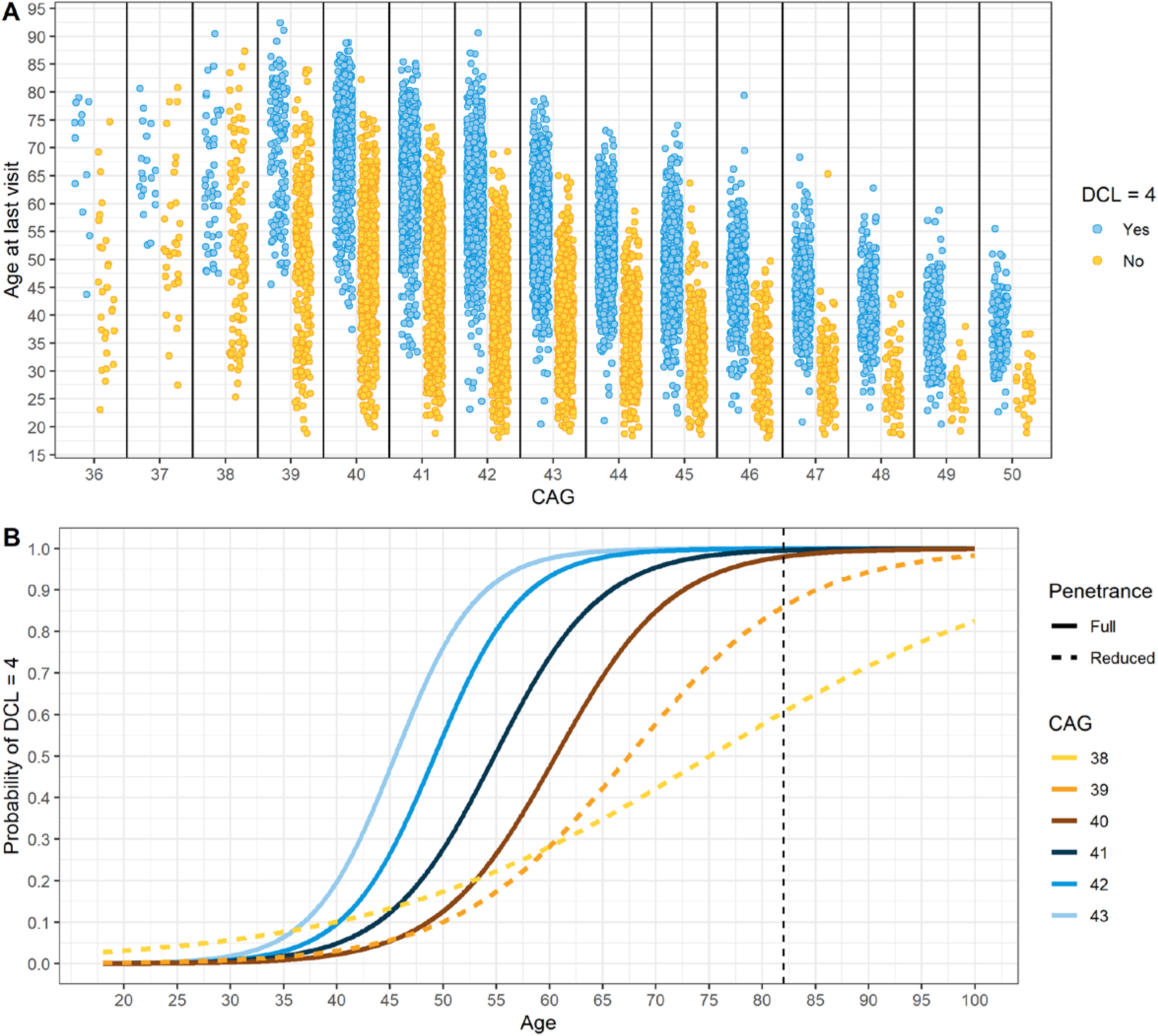
Reduced penetrance analysis. Panel A shows the age at last recorded study visit as a function of CAG expansion and rater assignment of DCL4 (Yes/No) for N = 12,152 participants from all the studies. Panel B shows curves from fitting a logistic mixed effects regression predicting DCL4 status (Yes/No) for each CAG expansion using the longitudinal data of each cohort. The vertical dashed line at 82 denotes the 2019 life expectancy of a western European.^42^

#### Is there reduced or age-dependent penetrance of HD in individuals with 36-39 CAGs?

Penetrance is defined as the proportion of individuals with a given genotype who exhibit the phenotype associated with that genotype.^41^ The phenotype was defined as reaching HD clinical motor diagnosis, here DCL4, in an expected life span (82 years).^42^ Allele lengths of 36-39 CAG are termed “reduced penetrance” implying that some, but not all, individuals in this population will exhibit the underlying pathology and overt signs of HD. It is possible that, just like for CAG ≥ 40, there is age-dependent penetrance, and with sufficient life span, all individuals with CAG = 36-39 would present with HD.^35-38,43-50^ We concluded that it is reduced penetrance in this CAG range based on robust evidence from population-based epidemiological studies and case reports,^45,46,48,49,51-53^ and our novel modeling analyses illustrated here (full statistical analysis and results in supplementary material).

Figure 1A shows that the proportion of individuals of ≥82 years that have not reached DCL4 decreases with increasing CAG length (38 CAG: 3/6, 39 CAG: 3/21, 40 CAG: 1/41, 41 CAG: 1/40 and 42 CAG: 0/10). Figure 1B shows that the predicted probabilities converge to 1 (complete penetrance) before the reference age of 82 for 41-43 CAG. For 40 CAG, the convergence to 1 is shortly after the reference age (86 years), and the curve looks similar to those of 41-43 CAG. In contrast, for 38-39 CAG, convergence is at >100 years old and the shape of the curves shift as CAG decreases (Table S6). Therefore, 36-39 CAG is necessary but not sufficient to define the disease state.

We adopted the following definition of HD:

***Huntington’s disease is defined by the presence of a CAG expansion in exon 1 of the HTT gene of either (a) CAG ≥ 40; or (b) CAG ≥ 36 and the presence of a disease-specific biomarker or disease-specific clinical syndrome***.

Because research is needed to establish a disease-specific biomarker or clinical syndrome with high specificity, high positive predictive value, and compelling biological plausibility to operationally define HD for CAG = 36-39, at this time, the staging will focus on individuals with CAG ≥ 40.

### HD Staging System

#### What is the temporal sequence of events in the natural history of HD?

Observational data and modeling analyses^26,54-57^ provide evidence that HD progression initiates with the genetic mutation, followed by the addition of measurable indicators of underlying pathophysiology (including neurodegeneration), then a detectable clinical sign or symptom, then a detectable functional change.

HD-ISS Stage 0 includes all individuals that carry a pathogenic CAG expansion but for whom there is no detectable change in pathological biomarkers, or signs or symptoms, or functional changes associated with HD. Stage 0 includes the process of neurodevelopment in the presence of the expanded *HTT* CAG tract and includes a window before pathogenesis or clinical presentation. The disease process is a continuum beginning *in utero*,^58,59^ resulting in the eventual presentation of an HD clinical phenotype impacting functional ability.

The pathogenesis that eventually initiates,^26,60^ and is then ongoing, can be detected by the measurable change in biomarkers indicating underlying neurodegeneration and signals Stage 1 and a change in prognosis. The presentation of definite clinical signs and symptoms, which can encompass cognitive or motor manifestations of HD, signals entry into Stage 2. Stage 3 is triggered by changes in an individual’s functional ability, such as a change in job status or difficulties performing activities of daily living.

In summary, this temporal sequence is conceptualized as follows:

**HD-ISS:**

***Stage 0: CAG ≥ 40***

***Stage 1: CAG ≥ 40 & biomarker of pathogenesis***

***Stage 2: CAG ≥ 40 & biomarker of pathogenesis & sign/symptom***

***Stage 3: CAG ≥ 40 & biomarker of pathogenesis & sign/symptom & functional change***

#### Which landmarks are best suited to classify HD individuals into each Stage?

Systematic literature review generated an exhaustive list of 2790 candidate assessments used in HD clinical research and we considered 16 potential landmarks (see Table S3) that had been evaluated in longitudinal studies of large sample size and are prognostic of further disease progression (Tables S1, S2, and S4).

#### Stage 1 Landmarks

We identified volumetric neuroimaging assessments for six regions as biological markers of pathogenesis. In natural history modeling, caudate vMRI and putamen vMRI, adjusted for head size, were consistently the earliest imaging biomarkers to show atrophy in HD and therefore we selected these to capture the transition into the process of neurodegeneration.

#### Stage 2 Landmarks

Candidates included motor and cognitive assessments but no behavioral ones; this is not unexpected since the latter typically do not progress with HD.^61-63^ To accommodate diversity in HD presentation, we selected one motor and one cognitive assessment as Stage 2 landmarks. To capture a more comprehensive motor construct, we selected the full Total Motor Score (TMS) over two of its components. In the cognitive domain, the Symbol Digit Modalities Test (SDMT) was the most widely reported in clinical research, while also demonstrating less combined impact of language and practice effects.^64,65^

#### Stage 3 Landmarks

We selected the Independence Scale (IS) and the Total Functional Capacity (TFC) over the TFC occupation sub-score because it captures broader information.

Given Stage 3’s decades-long duration, we further divided it into three broad conceptual groupings of mild, moderate, and severe functional deficits, defined to give clear boundaries without relying on additional specific quantitative landmarks:

> ***Mild***: *Individual does not require assistance with routine activities, though they may be difficult and/or take a long time*
>
> ***Moderate***: *Individual requires assistance with some routine activities*
>
> ***Severe***: *Individual cannot perform any routine activities independently*

The landmarks and Stage conditions selected are depicted in Figure 2.

**Figure 2.**
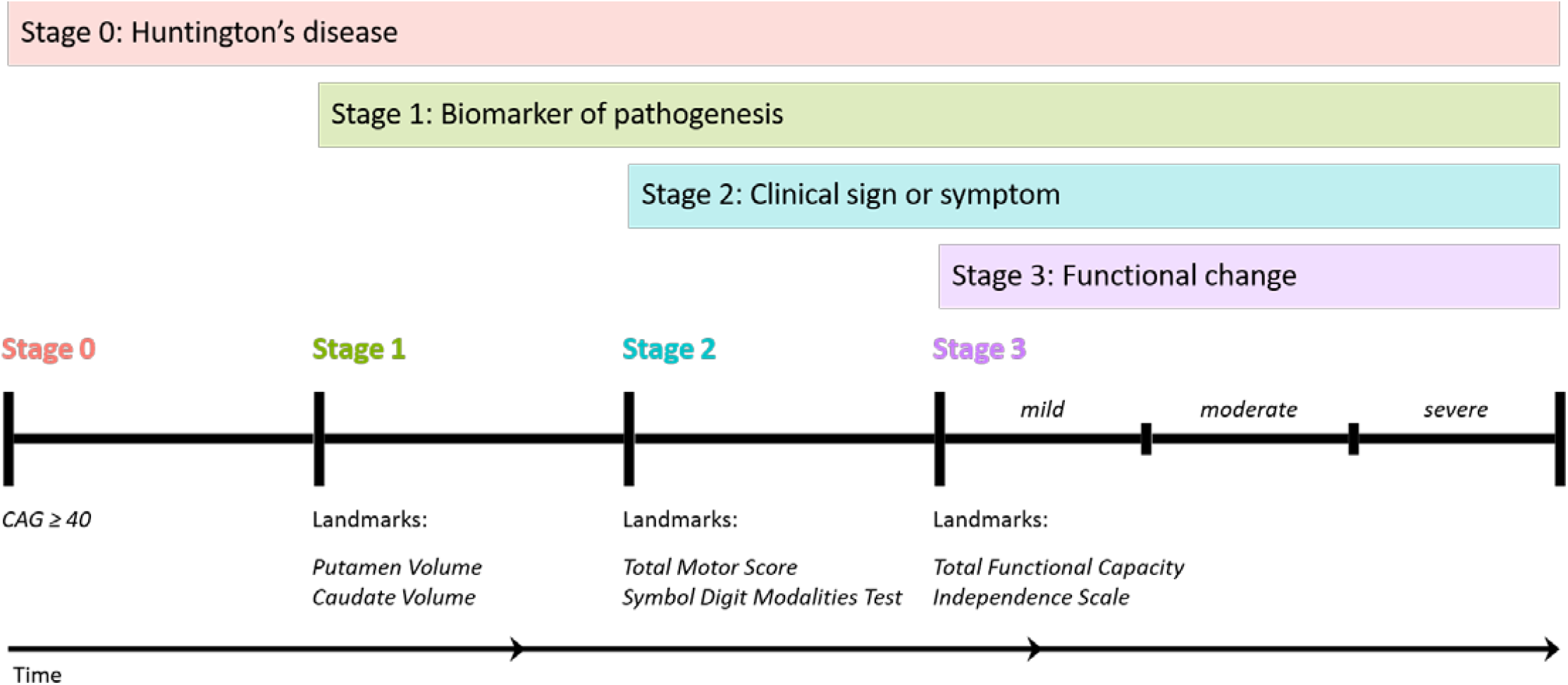
Cumulative Staging framework and landmarks. Graphical representation of the temporal sequence of Stage progression and the associated landmark assessments that define Stage entry. (Note: time not to scale)

#### What are the landmark cut-offs to operationalize the Stage criteria?

To determine cut-offs, we chose the quantile associated with the 5% tail area of the control population to define entry into each Stage and applied it to all of the landmark variables (either upper or lower tail areas, Figure S1).^66-68^ For SDMT we stratified the data into two groups based on education level.^69^ Importantly, because the cut-offs are based on control data they are independent of CAG length. Table 2 details an abbreviated cut-off look-up table aggregated by five-year intervals and Table S8 displays yearly cut-off values. As an alternative to the look-up tables, we have also created a Stage “calculator” available as an online tool (https://enroll-hd.org/calc/html_basic.htm). Given a person’s age and landmark assessment values, the calculator will output the Stage.

**Table 2.** Abridged landmark cut-off values by age. Putamen and caudate volumes are divided by intra-cranial volume and multiplied by 1000 (the table values can be divided by 1000 to produce the cut-offs as proportions).

| Age | Condition: Stage 1 |  | Condition: Stage 2 |  |  | Condition: Stage 3 |  |
| --- | --- | --- | --- | --- | --- | --- | --- |
|  | Putamen | Caudate | TMS | SDMT<sup>a</sup> | SDMT<sup>b</sup> | TFC | IS |
| 20 | 6.01 | 4.30 | 6 | 32 | 45 | 13 | 100 |
| 25 | 5.92 | 4.23 | 6 | 32 | 44 | 13 | 100 |
| 30 | 5.84 | 4.17 | 6 | 32 | 44 | 13 | 100 |
| 35 | 5.75 | 4.10 | 6 | 32 | 44 | 13 | 100 |
| 40 | 5.66 | 4.04 | 6 | 31 | 43 | 13 | 100 |
| 45 | 5.57 | 3.97 | 6 | 30 | 42 | 13 | 100 |
| 50 | 5.48 | 3.90 | 6 | 29 | 41 | 13 | 100 |
| 55 | 5.39 | 3.84 | 6 | 27 | 39 | 13 | 100 |
| 60 | 5.31 | 3.77 | 7 | 24 | 36 | 12 | 99 |
| 65 | 5.22 | 3.71 | 9 | 21 | 33 | 12 | 98 |
| 70 | 5.13 | 3.64 | 10 | 18 | 30 | 11 | 96 |
| 75 | 5.04 | 3.58 | 12 | 14 | 26 | 11 | 92 |
| 80 | 4.95 | 3.51 | 14 | 10 | 22 | 10 | 89 |
| 85 | 4.87 | 3.45 | 17 | 6 | 18 | 9 | 85 |
| 90 | 4.78 | 3.38 | 20 | 2 | 14 | 8 | 81 |

<sup>a</sup> Less than or equal to high school education
<sup>b</sup> Greater than high school education

### Internal Validation of the Staging System

#### Does Stage entry follow expected patterns?

Figure 3 shows the predicted probability of crossing a cut-off and fulfilling a Stage condition as a function of age for CAG = 42 (curves for other CAG are similar). The Stage condition curves are shifted to the left of the individual landmark curves for each domain and are time-ordered.

**Figure 3.**
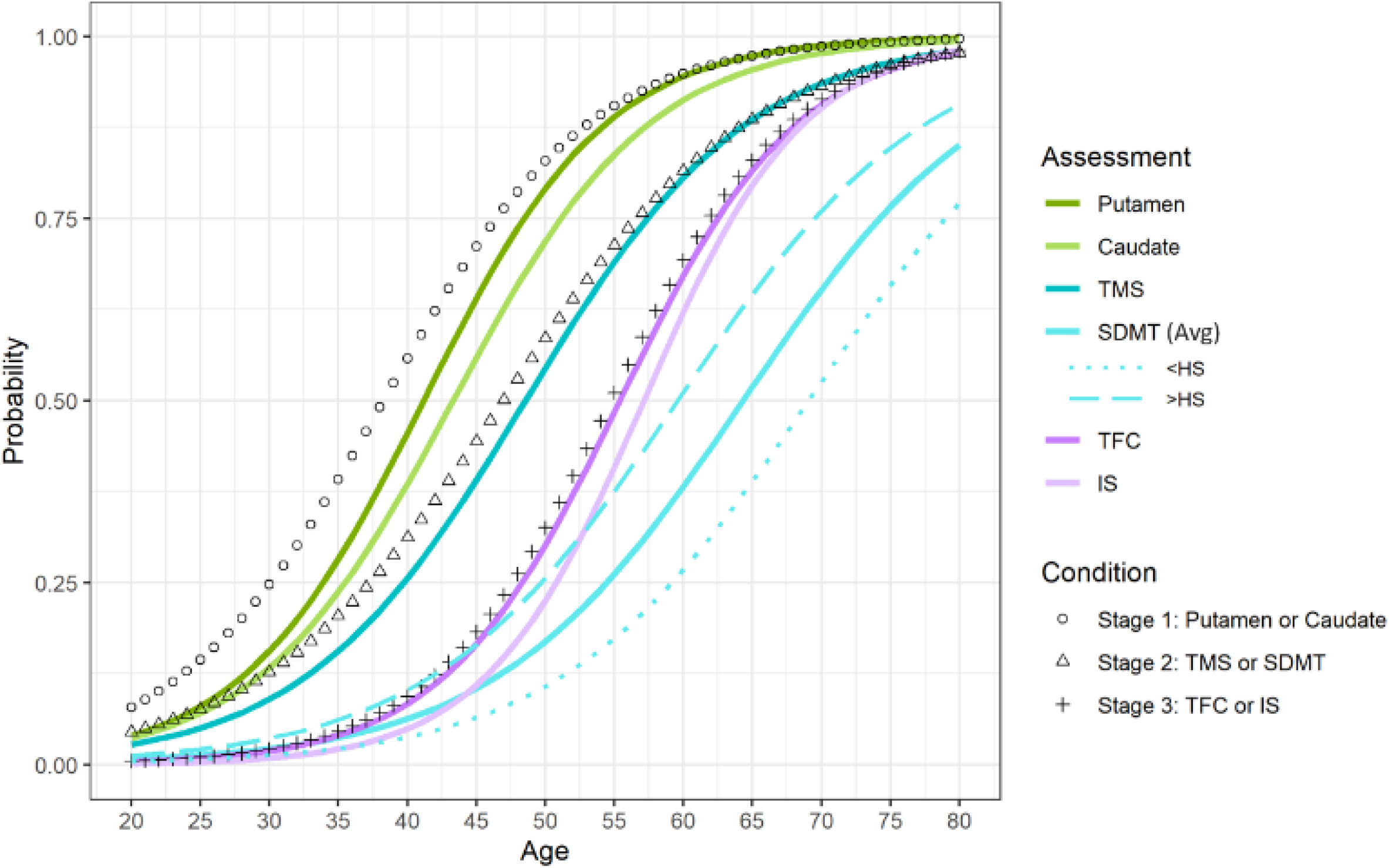
Assessment and Stage progression curves for CAG = 42. Probability of crossing the cut-off for an assessment (lines) and fulfilling the condition (shapes) as a function of age. The SDMT solid line depicts the average probability over education, the SDMT dotted line shows the probability for low education (high school education or less, <HS) and the SDMT dashed line shows the probability for high education (greater than high school education, >HS).

There is a similar sequential pattern for individual landmark curves. Note the SDMT curve crosses over the curves for the functional assessments because SDMT more gradually indexes HD progression (i.e., discriminates relative to controls at older ages). At younger ages the decline of SDMT for those with more education precedes the decline in TFC and IS, demonstrating that some individuals will meet the operational cut-off for SDMT before they show a functional change.

#### Does analysis of individual study visits reveal expected Staging patterns?

Evidence of ordered staging was assessed by scoring whether the individual met the cut-offs for each of the four Stages at that visit. A visit was only included in this analysis if there was complete data for all seven of the landmarks, including imaging data. Examination of Stage criteria patterns at each visit revealed that 87% of the patterns were consistent with the system (see Table S9).

#### Do individual participants progress through the Stages over time?

Overall, >85% of the participants had Kendall’s tau-a values^32^ that were consistent with orderly progression through the Stages; this percentage increased with more study visits (Table S10).

#### Are longitudinal models of group-level progress consistent with the Staging construct?

The estimated marginal (or in-Stage) probabilities and 95% CIs were computed from the estimated longitudinal continuation-ratio models (supplementary material). Figure 4 shows in-Stage probabilities by age and paneled by CAG length. In all panels the Stage 0 curve begins at a high probability because it is constitutive and starts at birth. This curve is always decreasing because there is only transition out of Stage 0. In contrast, Stage 3 is increasing because there is only transition into this Stage. The curves for Stage 1 and Stage 2 wax and wane because of transitions into and out of these Stages.

**Figure 4.**
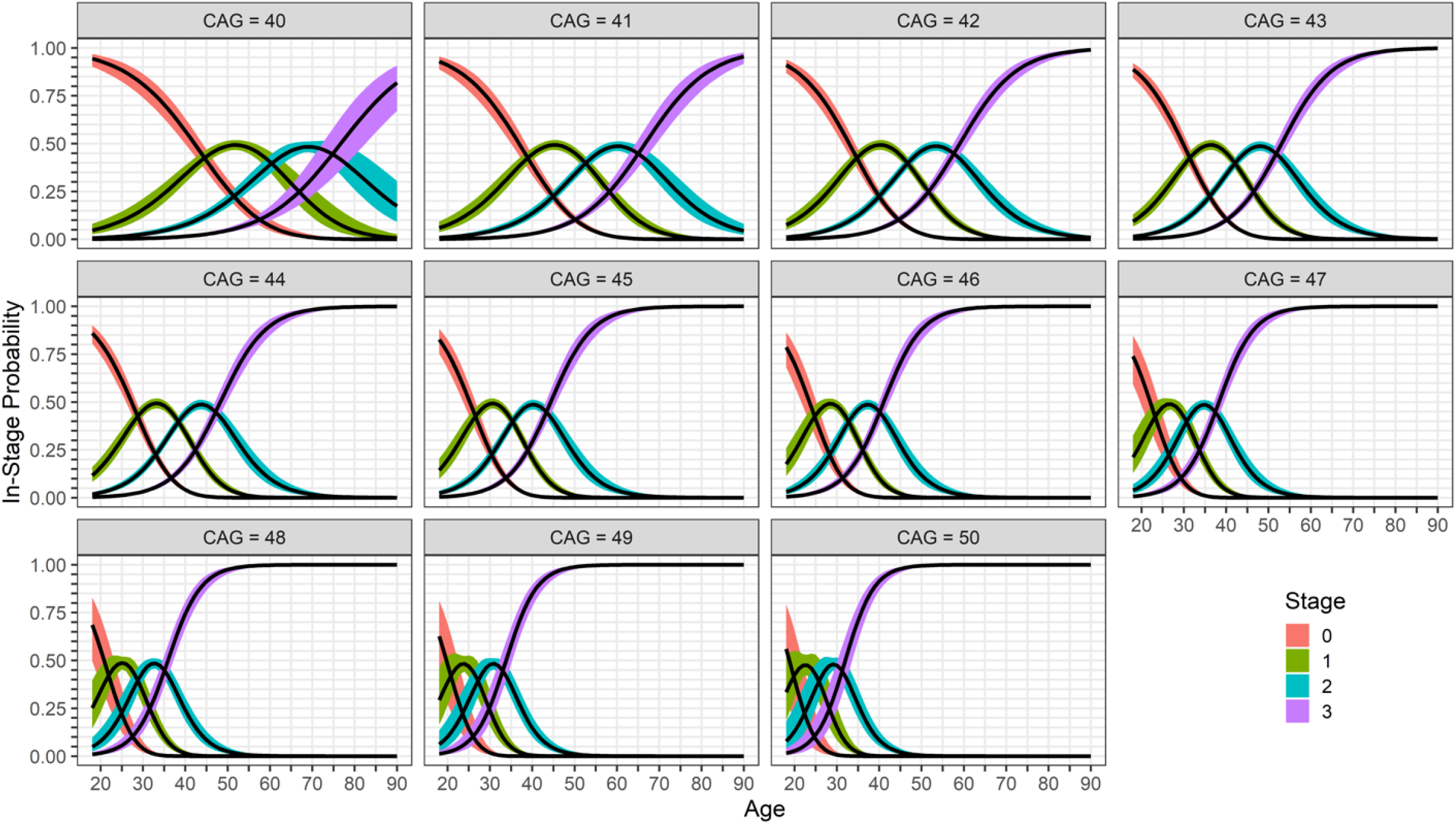
Stage probability by CAG. The probability of being in a Stage (or in-Stage probability) as a function of age with a colored ribbon representing the 95% confidence interval. Each panel represents a different CAG length.

CAG length effects are also evident when comparing panels in Figure 4; as CAG increase, the Stages still proceed in chronological sequence, but transitions shift to younger ages. The panels also show that the rates of transitions increase with CAG, such that it takes less time to progress through the Stages as CAG increase. Thus, not only do the Stage transitions occur at younger ages as CAG lengthen, but the transitions also happen faster, consistent with the known natural history of HD. Importantly, these probability curves are consistent with the HD-ISS framework for all CAG lengths evaluated.

Additional context for interpreting the HD-ISS Stages is provided by examining how they overlay with imaging and clinical variables over time (Figure 5, and supplementary Figure S5). For the cohort of 42 CAG, we fitted longitudinal models for the key clinical landmark assessments in order to predict the mean of each variable over time. Panel 5A shows stage probabilities and the predicted means transformed to a unit scale based on the proportional difference from controls.

**Figure 5.**
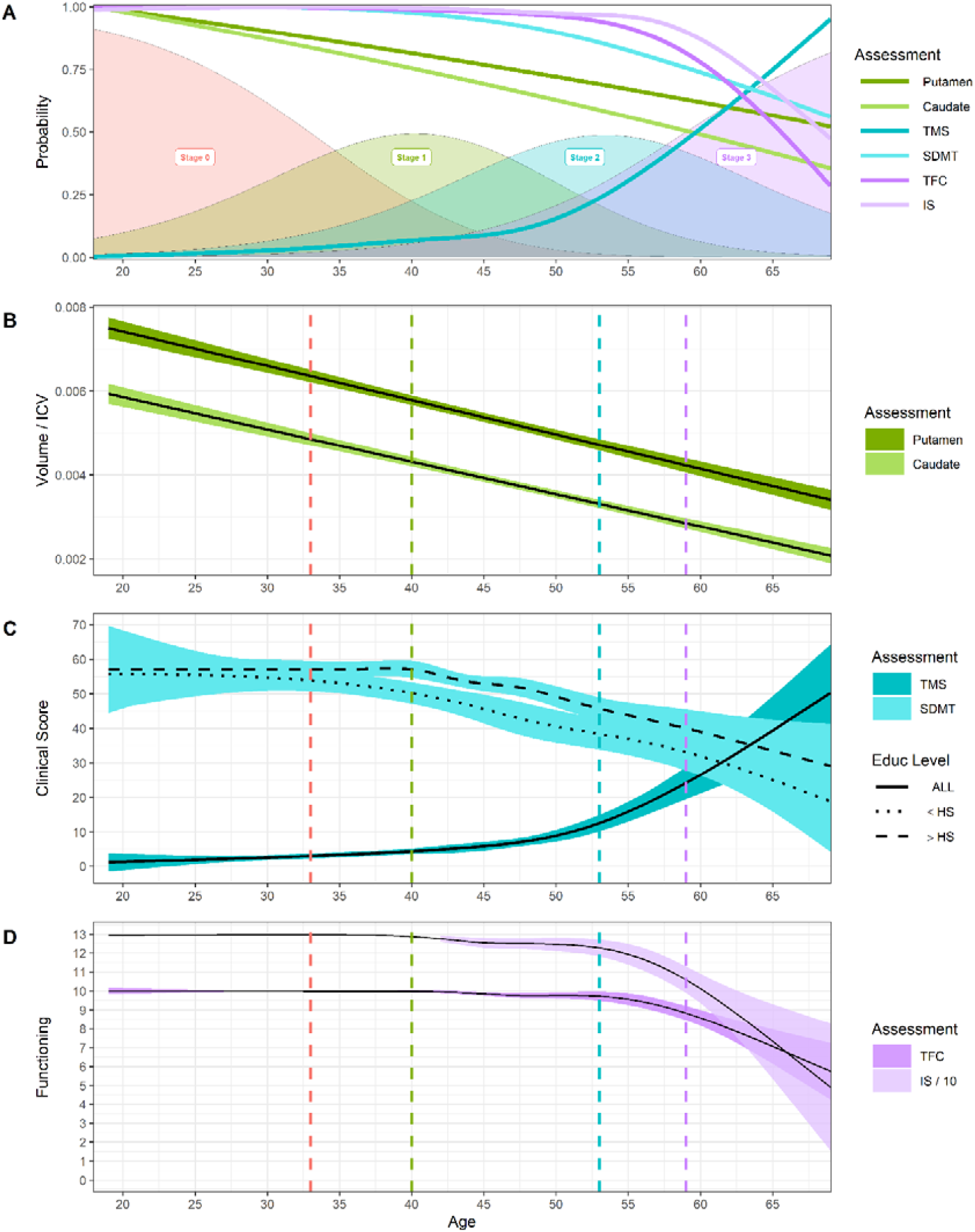
Overview of the HD-ISS for CAG = 42. Panel A shows the Stage probabilities (shaded regions) and the predicted change of the assessments as a proportion of the controls (which maps to a unit scale) as a function of age (see supplementary Figure S5). Panel B shows predicted mean putamen and caudate volumes divided by intra-cranial volume (ICV) with 95% CIs. Panel C shows predicted mean TMS and SDMT, with two education levels for SDMT (educ ≤ HS, educ > HS). Panel D shows the predicted mean TFC and predicted mean IS divided by 10. In Panels B-D a vertical dashed line (stage colors as in Panel A) denotes the age at which a Stage probability = 0.50.

Putamen and caudate volumes decline linearly over the Stage probabilities (Panel 5B), whereas the clinical variables have a non-linear trend in which an initial consistency eventually gives way to rapid decline (Panels 5C and 5D). TMS shows its fastest rate of change on the descending limb of Stage 2 and the ascending limb of Stage 3. There appears to be a similar effect for SDMT, though the maximum rate might occur slightly earlier. TFC and IS decline fastest later when the probability of being in Stage 3 is relatively high. The correspondence of the cUHDRS^70^ and DCL to the HD-ISS is seen in Figure S4. While entry into each HD-ISS Stage marks an inflection point in the progression of the landmark assessments, each continues to change as the disease progresses.

## DISCUSSION

We present a new paradigm to define and stage HD, primarily intended for use in HD clinical research. We establish a standard HD case definition based on *HTT* CAG expansion (≥40) and a formal staging system for HD, the HD-ISS, using well-established prognostic biological, clinical, and functional landmarks. We provide preliminary evidence that the HD-ISS is a valid system of classification. We show it matches the known natural history of HD in that the residence times in each stage are in line with the empirical expectation, the age at crossing a stage boundary is CAG-dependent as expected from previous research,^71-73^ and Stages are sequential, progressive, and align with age.

The HD-ISS used the great wealth of clinical and biomarker data available in accessible datasets to develop an integrated classification and staging system without reference to clinically-derived concepts such as “manifest,” “premanifest,” or “prodromal”; it also provides methodology for clinical study of HD before the emergence of a clinical syndrome and allows for testing of preventive interventions.^29,74^ Indeed, the biggest advantage of the HD-ISS is the ability to recognize, classify consistently, and follow-up with individuals very early in the course of the disease. This fills a clear void in HD research because the Shoulson and Fahn system does not classify individuals without obvious clinical signs or symptoms^12^; the more recent proposal from the MDS task force addressed earlier phases of the disease but remains incomplete because it lacks objective biomarker data and analysis of how these biomarkers behave in healthy controls, without which the full range of HD cannot be precisely characterized.^22^ Future work will establish a mapping between the HD-ISS and other systems, including Shoulson and Fahn.^75,76^

We followed formal consensus methodology and an evidence-based approach; nevertheless, there were some limitations. For example, the systematic literature review was restricted to English.^77,78^ Additionally, the imaging datasets, while highly cited in the literature, may suffer from selection bias because of their small size, geographic constraints, and required knowledge of genetic status. However, the key measures of interest (brain volumes) are quantitative and therefore less influenced by the study population’s demographics. Additionally, Enroll-HD, like any cohort, may suffer from selection bias; however, the database is robust because of its size and duration. The control participants analyzed share environmental exposures with the HD individuals and are similar to a larger normative population.^69^ We used the same datasets to establish cut-off values and validate Staging as we used to inform the conceptualization of the system, which could lead to overfitting; further research with additional data will allow for any necessary corrections.

Applying the HD-ISS in research will require additional considerations. First, while the implementation of vMRI requires specific technical considerations (supplementary material) and may be a hurdle,^79^ it would be essential for research studies to determine whether an individual is displaying neurodegeneration and is therefore in Stage 1. Second, our analysis indicates that the rate of “non-sequential individuals” in HD is similar to that in AD^66,80,81^; if there are HD individuals that biologically do not follow the sequential pattern, some may have a non-HD-related reason. Either way, they may be best excluded from studies. Third, while the HD-ISS is sufficiently flexible to incorporate new data, it is important for the system to maintain stability and comparability. Consequently, to establish a new landmark assessment, it will be important to understand how it affects classification accuracy and Stage boundaries. Finally, special considerations apply to either end of the HD genetic continuum. Although there is no maximum CAG in the HD definition, and cases of HD occur in children and adolescents (e.g. at CAG ≥ 56)^82^ and are included in the biological definition of HD, future research is needed to determine how the HD-ISS may apply specifically to this population. Individuals with reduced-penetrance alleles (36-39 CAG) would need to present with a disease-specific biomarker or disease-specific clinical syndrome to be classified as having HD in the HD-ISS. Practically, while this HD population should be given more attention in clinical research, the inclusion of individuals with reduced-penetrance alleles in interventional research would require careful consideration.

Even if only adopted for research, we recognize that this system affects HD families by acknowledging pathology prior to the current concept of clinical motor diagnosis. While this may be difficult, it will advance therapeutic development and may allow for expanded drug labeling aimed at preventing clinical decline. We are appreciative of the feedback from HD-COPE and look forward to future partnership with HD families.

In conclusion, we have developed a biological definition of HD and an evidence-based staging system of the entire disease course. The HD-ISS uses a numerical Staging that is similar to that adopted by the FDA Guidance for AD.^83^ We hope the HD research community will adopt this biological HD definition and the HD-ISS in all future clinical research studies. These Stages are not meant to be the sole inclusion criteria; more specific research participant selection, enrichment, and stratification approaches will be used to define subsets of the population that are particularly suitable for a given research project or therapeutic intervention. The HD-ISS could also be a biological paradigm for other genetic neurodegenerative diseases.^84^ This framework will enable the community to work together to bring hope to HD families and change the trajectory of HD.

### Future Directions

The HD-ISS encompasses the entire course of HD and will enable a common language for the standardization of research. Broadly sharing clinical study data will allow for evaluation of the HD-ISS’s performance in a prospective setting. We expect that these Stages have intrinsic clinical relevance because each represents a substantive difference in the pathophysiological state from the previous Stage; future research will be needed to evaluate the correspondence between the HD-ISS and existing measures, scales, and staging systems. Additional data on the reduced-penetrance allele population will allow the nomination and adoption of disease-specific biomarkers or clinical syndromes that meet the agreed-upon criteria.

The complementary work to the creation of the HD-ISS will be the analysis and nomination of enrichment biomarkers and outcome assessments appropriate for each Stage and across Stages. By discussing trial design and endpoints, we hope to advance regulatory science in support of drug development across the spectrum of HD. In the future, we anticipate that the HD-ISS will contribute to informing the development of practical regulatory strategies for disease-modifying (and potentially preventative) HD treatments.

## RESEARCH IN CONTEXT

### Evidence before this study

Huntington’s disease (HD) clinical research and case definitions to date have been structured around clinical motor diagnosis. However, the clinical definition of HD was conceptualized before the full understanding of either HD’s genetic cause or the pathobiology that precedes HD clinical signs and symptoms. The existing terminology is focused on this relatively advanced “manifest” phase of disease and is not standardized or based on biology. Currently, only one staging system is widely used in HD, but it is only applicable to advanced disease. Until now, HD clinical research studies that include participants before clinical motor diagnosis have developed distinct methods to select and categorize participants. Recently, a Movement Disorder Society task force proposed terminology to standardize these populations based on clinical criteria, but without the incorporation of biomarker data, it is difficult to precisely characterize early disease progression. Therefore, the HD research community needs a system to describe the full course of disease from HD inception (birth) to death. This work was inspired by the precedent led by NIA-AA and the solution adopted in the FDA guidance for Alzheimer’s disease.

### Added value of this study

For the first time, the HD-ISS establishes a biological definition of HD that considers the spectrum of known CAG expansions and an evidence-based staging system that allows for the research classification of HD individuals in any phase of the disease. Importantly, the HD-ISS addresses the decades-long gap in disease course before clinical signs and symptoms are unequivocal, providing the ability to identify and consistently classify individuals from birth.

Furthermore, the HD-ISS affords common language and terminology to facilitate future data sharing and merging of datasets. Additionally, the Stage boundary landmark assessment cut-offs are based on empirical analysis of the difference between HD individuals and controls. These cut-offs are the first in the field and provide an important novel tool.

### Implications of all the available evidence

This HD framework provides a new structure for the conduct of clinical studies, representing a needed shift to allow for the possibility of testing interventions that prevent or delay the initial onset of symptoms and take place before current HD clinical motor diagnosis. By harmonizing both concepts and language, the HD-ISS can be immediately applied to enable standardized definitions of study populations and to facilitate cross-study comparison of inclusion criteria. Further, the HD-ISS will be an important foundation on which to build future regulatory strategies and guidance on appropriate outcome assessments for study participants in a specific Stage and across Stages.

## Data Availability

We used four publicly available data sets: Enroll-HD (4th periodic data set), IMAGE-HD, PREDICT-HD, and TRACK-HD. Contact for more information regarding data access.

https://enroll-hd.org/

## ACKNOWLEDGEMENTS

The authors wish to thank HD-COPE for their helpful input. Data used in this work was generously provided by the participants in the Enroll-HD study and made available by CHDI Foundation, Inc. Enroll-HD is a global clinical research platform intended to accelerate progress towards therapeutics for Huntington’s disease; core datasets are collected annually on all research participants as part of this multi-center longitudinal observational study. Enroll-HD is sponsored by CHDI Foundation, Inc., a nonprofit biomedical research organization exclusively dedicated to developing therapeutics for Huntington’s disease. Enroll-HD would not be possible without the vital contribution of the research participants and their families. We would like to acknowledge Monash University (Clayton, Australia) for the use of the IMAGE-HD data (project led by principal investigator Professor Nellie Georgiou-Karistianis). Data used in this work was generously provided by the participants in PREDICT-HD and made available by the PREDICT-HD Investigators and Coordinators of the Huntington Study Group, Jane Paulsen, Principal Investigator. PREDICT-HD was funded by the National Institute of Health (NIH) under Grant# NS040068. Data used in this work was generously provided by the participants in the TRACK-HD study and made available by Dr. Sarah Tabrizi, Principal Investigator, University College London. The authors also extend their gratitude to the study participants and their families who supported them. We thank Robi Blumenstein, Martha Brumfield, Darren Freeman, Simon Noble, Chris Rackow, and Swati Sathe for their input and support during this project and in manuscript drafting. We thank Teresa Moogan for her copyediting skills. We appreciate the support of the HD-RSC Coordinating Committee and their useful feedback, and thank Danielle Gartner, Jacqueline Major, and Sandra Gonzalez for administrative support of consortium activities.

## AUTHOR CONTRIBUTIONS

SJT and SS co-chaired the working group. ECG and AM contributed to the concept of the statistical analysis and design of the figures, writing original drafts and several versions of both the report to the HD-RSC coordinating committee and the manuscript, and managed manuscript revisions. SJT, SS, CS, and JDL contributed to the first drafts of the report and the manuscript. APM contributed to the setup and organization of the project. SJT, SS, BB, PK, TAM, JP, CAR, MZ, JDL, and CS voted on all consensus decisions. JDL developed the concept of the statistical analyses and conducted formal analysis and validation. CS led the design of the project and the methodology. All authors participated in working group meetings, contributed to the various drafts of the report and the manuscript, and reviewed and revised the final draft of the manuscript.

## REFERENCES

1 Bates, G. P. et al. Huntington disease. Nat Rev Dis Primers 1, 15005, doi:10.1038/nrdp.2015.5 (2015).

2 Oosterloo, M. et al. Disease Onset in Huntington’s Disease: When Is the Conversion? Mov Disord Clin Pract 8, 352–360, doi:10.1002/mdc3.13148 (2021).

3 McCusker, E. A. & Loy, C. T. Huntington Disease: The Complexities of Making and Disclosing a Clinical Diagnosis After Premanifest Genetic Testing. Tremor Other Hyperkinet Mov (N Y) 7, 467, doi:10.7916/D8PK0TDD (2017).

4 Ross, C. A. et al. Huntington disease: natural history, biomarkers and prospects for therapeutics. Nat Rev Neurol 10, 204–216, doi:10.1038/nrneurol.2014.24 (2014).

5 Kim, S. D. & Fung, V. S. An update on Huntington’s disease: from the gene to the clinic. Curr Opin Neurol 27, 477–483, doi:10.1097/WCO.0000000000000116 (2014).

6 Reilmann, R., Leavitt, B. R. & Ross, C. A. Diagnostic criteria for Huntington’s disease based on natural history. Mov Disord 29, 1335–1341, doi:10.1002/mds.26011 (2014).

7 Tabrizi, S. J. et al. Predictors of phenotypic progression and disease onset in premanifest and early-stage Huntington’s disease in the TRACK-HD study: analysis of 36-month observational data. Lancet Neurol 12, 637–649, doi:10.1016/S1474-4422(13)70088-7 (2013).

8 Gusella, J. F. et al. A polymorphic DNA marker genetically linked to Huntington’s disease. Nature 306, 234–238, doi:10.1038/306234a0 (1983).

9 A novel gene containing a trinucleotide repeat that is expanded and unstable on Huntington’s disease chromosomes. The Huntington’s Disease Collaborative Research Group. Cell 72, 971–983, doi:10.1016/0092-8674(93)90585-e (1993).

10 Hogarth, P. et al. Interrater agreement in the assessment of motor manifestations of Huntington’s disease. Mov Disord 20, 293–297, doi:10.1002/mds.20332 (2005).

11 Liu, D. et al. Motor onset and diagnosis in Huntington disease using the diagnostic confidence level. J Neurol 262, 2691–2698, doi:10.1007/s00415-015-7900-7 (2015).

12 Shoulson, I. & Fahn, S. Huntington disease: clinical care and evaluation. Neurology 29, 1–3, doi:10.1212/wnl.29.1.1 (1979).

13 Tabrizi, S. J. et al. Biological and clinical manifestations of Huntington’s disease in the longitudinal TRACK-HD study: cross-sectional analysis of baseline data. Lancet Neurol 8, 791–801, doi:10.1016/S1474-4422(09)70170-X (2009).

14 Paulsen, J. S. et al. Preparing for preventive clinical trials: the Predict-HD study. Arch Neurol 63, 883–890, doi:10.1001/archneur.63.6.883 (2006).

15 Huntington Study Group, C. I. & Dorsey, E. Characterization of a large group of individuals with huntington disease and their relatives enrolled in the COHORT study. PLoS One 7, e29522, doi:10.1371/journal.pone.0029522 (2012).

16 Huntington Study Group, P. I. At risk for Huntington disease: The PHAROS (Prospective Huntington At Risk Observational Study) cohort enrolled. Arch Neurol 63, 991–996, doi:10.1001/archneur.63.7.991 (2006).

17 Orth, M. et al. Observing Huntington’s Disease: the European Huntington’s Disease Network’s REGISTRY. PLoS Curr 2, RRN1184, doi:10.1371/currents.RRN1184 (2010).

18 Landwehrmeyer, G. B. et al. Data Analytics from Enroll-HD, a Global Clinical Research Platform for Huntington’s Disease. Mov Disord Clin Pract 4, 212–224, doi:10.1002/mdc3.12388 (2017).

19 Wilkes, F. A. et al. Striatal morphology and neurocognitive dysfunction in Huntington disease: The IMAGE-HD study. Psychiatry Res Neuroimaging 291, 1–8, doi:10.1016/j.pscychresns.2019.07.003 (2019).

20 Scahill, R. I. et al. Biological and clinical characteristics of gene carriers far from predicted onset in the Huntington’s disease Young Adult Study (HD-YAS): a cross-sectional analysis. Lancet Neurol 19, 502–512, doi:10.1016/S1474-4422(20)30143-5 (2020).

21 Kloppel, S. et al. Compensation in Preclinical Huntington’s Disease: Evidence From the Track-On HD Study. EBioMedicine 2, 1420–1429, doi:10.1016/j.ebiom.2015.08.002 (2015).

22 Ross, C. A. et al. Movement Disorder Society Task Force Viewpoint: Huntington’s Disease Diagnostic Categories. Mov Disord Clin Pract 6, 541–546, doi:10.1002/mdc3.12808 (2019).

23 Carroll, J. New Collaboration Seeks to Speed Huntington’s Disease Drug Licensing., <https://en.hdbuzz.net/259.> (2018).

24 Nair, R., Aggarwal, R. & Khanna, D. Methods of formal consensus in classification/diagnostic criteria and guideline development. Semin Arthritis Rheum 41, 95–105, doi:10.1016/j.semarthrit.2010.12.001 (2011).

25 Georgiou-Karistianis, N. et al. Functional and connectivity changes during working memory in Huntington’s disease: 18 month longitudinal data from the IMAGE-HD study. Brain Cogn 83, 80–91, doi:10.1016/j.bandc.2013.07.004 (2013).

26 Georgiou-Karistianis, N., Scahill, R., Tabrizi, S. J., Squitieri, F. & Aylward, E. Structural MRI in Huntington’s disease and recommendations for its potential use in clinical trials. Neurosci Biobehav Rev 37, 480–490, doi:10.1016/j.neubiorev.2013.01.022 (2013).

27 Long, J. D., Paulsen, J. S., Investigators, P.-H. & Coordinators of the Huntington Study, G. Multivariate prediction of motor diagnosis in Huntington’s disease: 12 years of PREDICT-HD. Mov Disord 30, 1664–1672, doi:10.1002/mds.26364 (2015).

28 Paulsen, J. S. et al. Detection of Huntington’s disease decades before diagnosis: the Predict-HD study. J Neurol Neurosurg Psychiatry 79, 874–880, doi:10.1136/jnnp.2007.128728 (2008).

29 Paulsen, J. S. et al. Clinical and Biomarker Changes in Premanifest Huntington Disease Show Trial Feasibility: A Decade of the PREDICT-HD Study. Front Aging Neurosci 6, 78, doi:10.3389/fnagi.2014.00078 (2014).

30 Gonnella, J. S., Goran, M. J., Williamson, J. W. & Cotsonas, N. J., Jr. Evaluation of patient care. An approach. JAMA 214, 2040–2043 (1970).

31 Cebul, R. D., Hershey, J. C. & Williams, S. V. Using multiple tests: series and parallel approaches. Clin Lab Med 2, 871–890 (1982).

32 Kendall, M. G., and Gibbons, J. D.. Rank Correlation Methods. 5th edn, (Oxford, 1990).

33 Dos Santos, D. M. & Berridge, D. M. A continuation ratio random effects model for repeated ordinal responses. Stat Med 19, 3377–3388, doi:10.1002/1097-0258(20001230)19:24<3377::aid-sim526>3.0.co;2-e (2000).

34 Schomaker, M. & Heumann, C. Bootstrap inference when using multiple imputation. Stat Med 37, 2252–2266, doi:10.1002/sim.7654 (2018).

35 Wexler, N. S. et al. Venezuelan kindreds reveal that genetic and environmental factors modulate Huntington’s disease age of onset. Proc Natl Acad Sci U S A 101, 3498–3503, doi:10.1073/pnas.0308679101 (2004).

36 Pericak-Vance, M. A. et al. Genetic linkage studies in Huntington disease. Cytogenet Cell Genet 22, 640–645, doi:10.1159/000131042 (1978).

37 Duyao, M. et al. Trinucleotide repeat length instability and age of onset in Huntington’s disease. Nat Genet 4, 387–392, doi:10.1038/ng0893-387 (1993).

38 Laird, C. D. Proposed genetic basis of Huntington’s disease. Trends Genet 6, 242–247, doi:10.1016/0168-9525(90)90206-l (1990).

39 Brinkman, R. R., Mezei, M. M., Theilmann, J., Almqvist, E. & Hayden, M. R. The likelihood of being affected with Huntington disease by a particular age, for a specific CAG size. Am J Hum Genet 60, 1202–1210 (1997).

40 Stevens, D. Huntington Chorea: demography, clinics, genetics. MD thesis, University of London, (1976).

41 Griffiths, A. J. F., J. H. Miller, D. T. Suzuki, R. C. Lewontin, and W. M. Gelbart.. An Introduction to Genetic Analysis. 7th Edition edn, (W. H. Freeman, 2000).

42 WHO. Methods and data sources for life tables 1990-2019 (December 2020).

43 Findlay Black, H., Wright, G.E.B., Collins, J.A. et al. Frequency of the loss of CAA interruption in the HTT CAG tract and implications for Huntington disease in the reduced penetrance range.. Genet Med 22, 2108–2113 (2020).

44 Wright, G. E. B. et al. Interrupting sequence variants and age of onset in Huntington’s disease: clinical implications and emerging therapies. Lancet Neurol 19, 930–939, doi:10.1016/S1474-4422(20)30343-4 (2020).

45 Wright, G. E. B. et al. Length of Uninterrupted CAG, Independent of Polyglutamine Size, Results in Increased Somatic Instability, Hastening Onset of Huntington Disease. Am J Hum Genet 104, 1116–1126, doi:10.1016/j.ajhg.2019.04.007 (2019).

46 Rubinsztein, D. C. et al. Phenotypic characterization of individuals with 30-40 CAG repeats in the Huntington disease (HD) gene reveals HD cases with 36 repeats and apparently normal elderly individuals with 36-39 repeats. Am J Hum Genet 59, 16–22 (1996).

47 Cooper, D. N., Krawczak, M., Polychronakos, C., Tyler-Smith, C. & Kehrer-Sawatzki, H. Where genotype is not predictive of phenotype: towards an understanding of the molecular basis of reduced penetrance in human inherited disease. Hum Genet 132, 1077–1130, doi:10.1007/s00439-013-1331-2 (2013).

48 Quarrell, O. W. et al. Reduced penetrance alleles for Huntington’s disease: a multi-centre direct observational study. J Med Genet 44, e68, doi:10.1136/jmg.2006.045120 (2007).

49 McNeil, S. M. et al. Reduced penetrance of the Huntington’s disease mutation. Hum Mol Genet 6, 775–779, doi:10.1093/hmg/6.5.775 (1997).

50 Semaka, A., Creighton, S., Warby, S. & Hayden, M. R. Predictive testing for Huntington disease: interpretation and significance of intermediate alleles. Clin Genet 70, 283–294, doi:10.1111/j.1399-0004.2006.00668.x (2006).

51 Kay, C. et al. Huntington disease reduced penetrance alleles occur at high frequency in the general population. Neurology 87, 282–288, doi:10.1212/WNL.0000000000002858 (2016).

52 Langbehn, D. R. et al. A new model for prediction of the age of onset and penetrance for Huntington’s disease based on CAG length. Clin Genet 65, 267–277, doi:10.1111/j.1399-0004.2004.00241.x (2004).

53 Genetic Modifiers of Huntington’s Disease Consortium. Electronic address, g. h. m. h. e. & Genetic Modifiers of Huntington’s Disease, C. CAG Repeat Not Polyglutamine Length Determines Timing of Huntington’s Disease Onset. Cell 178, 887–900 e814, doi:10.1016/j.cell.2019.06.036 (2019).

54 Wijeratne, P. A. et al. An image-based model of brain volume biomarker changes in Huntington’s disease. Ann Clin Transl Neurol 5, 570–582, doi:10.1002/acn3.558 (2018).

55 Aziz, N. A., van der Burg, J. M. M., Tabrizi, S. J. & Landwehrmeyer, G. B. Overlap between age-at-onset and disease-progression determinants in Huntington disease. Neurology 90, e2099–e2106, doi:10.1212/WNL.0000000000005690 (2018).

56 Wild, E. J. et al. Rate and acceleration of whole-brain atrophy in premanifest and early Huntington’s disease. Mov Disord 25, 888–895, doi:10.1002/mds.22969 (2010).

57 Tang, C. & Feigin, A. Monitoring Huntington’s disease progression through preclinical and early stages. Neurodegener Dis Manag 2, 421–435, doi:10.2217/nmt.12.34 (2012).

58 Barnat, M. et al. Huntington’s disease alters human neurodevelopment. Science 369, 787–793, doi:10.1126/science.aax3338 (2020).

59 Nopoulos, P. C. et al. Smaller intracranial volume in prodromal Huntington’s disease: evidence for abnormal neurodevelopment. Brain 134, 137–142, doi:10.1093/brain/awq280 (2011).

60 Aylward, E. H. et al. Onset and rate of striatal atrophy in preclinical Huntington disease. Neurology 63, 66–72, doi:10.1212/01.wnl.0000132965.14653.d1 (2004).

61 Morkl, S. et al. Problem solving, impulse control and planning in patients with early-and late-stage Huntington’s disease. Eur Arch Psychiatry Clin Neurosci 266, 663–671, doi:10.1007/s00406-016-0707-4 (2016).

62 Simpson, J. et al. Validity of irritability in Huntington’s disease: A scoping review. Cortex 120, 353–374, doi:10.1016/j.cortex.2019.06.012 (2019).

63 Martinez-Horta, S. et al. Structural and metabolic brain correlates of apathy in Huntington’s disease. Mov Disord 33, 1151–1159, doi:10.1002/mds.27395 (2018).

64 Braisch, U. et al. Identification of symbol digit modality test score extremes in Huntington’s disease. Am J Med Genet B Neuropsychiatr Genet 180, 232–245, doi:10.1002/ajmg.b.32719 (2019).

65 Stout, J. C. et al. HD-CAB: a cognitive assessment battery for clinical trials in Huntington’s disease 1,2,3. Mov Disord 29, 1281–1288, doi:10.1002/mds.25964 (2014).

66 Koscik, R. L. et al. Longitudinal Standards for Mid-life Cognitive Performance: Identifying Abnormal Within-Person Changes in the Wisconsin Registry for Alzheimer’s Prevention. J Int Neuropsychol Soc 25, 1–14, doi:10.1017/S1355617718000929 (2019).

67 Clark, L. R. et al. Mild Cognitive Impairment in Late Middle Age in the Wisconsin Registry for Alzheimer’s Prevention Study: Prevalence and Characteristics Using Robust and Standard Neuropsychological Normative Data. Arch Clin Neuropsychol 31, 675–688, doi:10.1093/arclin/acw024 (2016).

68 Litvan, I. et al. Diagnostic criteria for mild cognitive impairment in Parkinson’s disease: Movement Disorder Society Task Force guidelines. Mov Disord 27, 349–356, doi:10.1002/mds.24893 (2012).

69 Mills, J. A., Long, J. D., Mohan, A., Ware, J. J. & Sampaio, C. Cognitive and Motor Norms for Huntington’s Disease. Arch Clin Neuropsychol 35, 671–682, doi:10.1093/arclin/acaa026 (2020).

70 Schobel, S. A. et al. Motor, cognitive, and functional declines contribute to a single progressive factor in early HD. Neurology 89, 2495–2502, doi:10.1212/WNL.0000000000004743 (2017).

71 Langbehn, D. R. et al. Association of CAG Repeats With Long-term Progression in Huntington Disease. JAMA Neurol, doi:10.1001/jamaneurol.2019.2368 (2019).

72 Podvin, S., Reardon, H. T., Yin, K., Mosier, C. & Hook, V. Multiple clinical features of Huntington’s disease correlate with mutant HTT gene CAG repeat lengths and neurodegeneration. J Neurol 266, 551–564, doi:10.1007/s00415-018-8940-6 (2019).

73 Lee, J. M. et al. CAG repeat expansion in Huntington disease determines age at onset in a fully dominant fashion. Neurology 78, 690–695, doi:10.1212/WNL.0b013e318249f683 (2012).

74 Langbehn, D. R. & Hersch, S. Clinical Outcomes and Selection Criteria for Prodromal Huntington’s Disease Trials. Mov Disord, doi:10.1002/mds.28222 (2020).

75 Ghosh, S. et al. An Exploration of Latent Structure in Observational Huntington’s Disease Studies. AMIA Jt Summits Transl Sci Proc 2017, 92–102 (2017).

76 Sun, Z. et al. A probabilistic disease progression modeling approach and its application to integrated Huntington’s disease observational data. JAMIA Open 2, 123–130, doi:10.1093/jamiaopen/ooy060 (2019).

77 Mueller, M. et al. Methods to systematically review and meta-analyse observational studies: a systematic scoping review of recommendations. BMC Med Res Methodol 18, 44, doi:10.1186/s12874-018-0495-9 (2018).

78 Morrison, A. et al. The effect of English-language restriction on systematic review-based meta-analyses: a systematic review of empirical studies. Int J Technol Assess Health Care 28, 138–144, doi:10.1017/S0266462312000086 (2012).

79 Jack, C. R., Jr. et al. The Alzheimer’s Disease Neuroimaging Initiative (ADNI): MRI methods. J Magn Reson Imaging 27, 685–691, doi:10.1002/jmri.21049 (2008).

80 Wei, Y., Pere, A., Koenker, R. & He, X. Quantile regression methods for reference growth charts. Stat Med 25, 1369–1382, doi:10.1002/sim.2271 (2006).

81 Ounpraseuth, S. T., Magann, E. F., Spencer, H. J., Rabie, N. Z. & Sandlin, A. T. Normal amniotic fluid volume across gestation: Comparison of statistical approaches in 1190 normal amniotic fluid volumes. J Obstet Gynaecol Res 43, 1122–1131, doi:10.1111/jog.13332 (2017).

82 Cronin, T., Rosser, A. & Massey, T. Clinical Presentation and Features of Juvenile-Onset Huntington’s Disease: A Systematic Review. J Huntingtons Dis 8, 171–179, doi:10.3233/JHD-180339 (2019).

83 FDA. Alzheimer’s Disease: Developing Drugs for Treatment Guidance for Industy, <https://www.fda.gov/regulatory-information/search-fda-guidance-documents/alzheimers-disease-developing-drugs-treatment-guidance-industy> (2018).

84 Katsuno, M., Sahashi, K., Iguchi, Y. & Hashizume, A. Preclinical progression of neurodegenerative diseases. Nagoya J Med Sci 80, 289–298, doi:10.18999/nagjms.80.3.289 (2018).

