## Supplementary Material for "Huntington’s Disease Integrated Staging System (HD-ISS): A Novel Evidence-Based Classification System For Staging"

Table of Contents


1. Organizational Details ... 2
2. RSF Meeting Record ... 3
3. Selection of Landmarks ... 4
4. Statistical Methods ... 16
   - A. Reduced Penetrance ... 16
     - i. Descriptive Statistics ... 16
     - ii. Inferential: Logistic Regression for Longitudinal Data ... 16
   - B. Participants for Staging System Development and Validation ... 17
   - C. Cut-off Values ... 19
     - i. Quantile Regression for Longitudinal Data ... 20
   - D. Scoring and Stage Assignment ... 21
   - E. Tail Area Justification ... 22
   - F. Missing Data ... 23
   - G. Stage Characteristics ... 25
     - i. Individual Visits ... 26
     - ii. Longitudinal Progression ... 26
   - H. Stage Probabilities ... 27
     - i. Continuation Ratio Regression for Longitudinal Data ... 27
   - I. Mapping Clinical Variables to Stage Probabilities ... 32
5. vMRI Technical Details ... 34
6. Supplementary Figure S5 ... 35
7. References ... 36

### 1. Organizational Details

- **Critical Path Institute (C-Path)**: an independent, nonprofit organization established in 2005 as a public and private partnership. C-Path's mission is to catalyze the development of new approaches that advance medical innovation and regulatory science, accelerating the path to a healthier world. An international leader in forming collaborations, C-Path has established numerous global consortia that currently include more than 1,600 scientists from government and regulatory agencies, academia, patient organizations, disease foundations, and dozens of pharmaceutical and biotech companies.

- **Huntington's Disease Regulatory Science Consortium (HD-RSC)**: an initiative created in a partnership between C-Path and CHDI Foundation, the HD-RSC was formally launched in March 2018 with the primary goal of creating new tools and methods to advance efficient clinical development and address the regulatory needs for approval of HD therapeutics**.** The HD-RSC provides the forum and structure to bring together necessary participants from the HD community for data contribution and tool development, leading to efficiencies in the development of new therapies. The HD-RSC is comprised of participants from government, academia, industry, non-profit research, and patient advocacy organizations.

- **Regulatory Science Forum Working Group (RSF)**: an HD-RSC working group responsible for developing white papers and/or publications based on HD community input to outline key points to consider for enabling clinical trials and development of therapeutics targeting HD individuals before clinical motor diagnosis.

- **CHDI Foundation, Inc.**: a privately funded nonprofit biomedical research organization exclusively dedicated to collaboratively developing therapeutics that will substantially improve the lives of those affected by HD. Our scientists work closely with a network of more than 700 researchers in academic and industrial laboratories around the world in the pursuit of these novel therapies, providing strategic scientific direction to ensure that our common goals remain in focus. This helps bridge the translational gap that often exists between academic and industrial research pursuits and that adds costly delays to therapy development. In its role as a collaborative enabler, CHDI seeks to bring the right partners together to identify and address critical scientific issues and move drug candidates to clinical evaluation as rapidly as possible. Our activities extend from exploratory biology to the identification and validation of therapeutic targets, and from drug discovery and development to clinical studies and trials. More information about CHDI can be found at www.chdifoundation.org.

- **Rancho Biosciences:** an independent research organization that provided data curation services for the RSF. Rancho Biosciences performs data searching and analysis using algorithmic methods that include natural language processing in order to organize, catalog, curate, and standardize a variety of unstructured and disorganized data including scientific literature, clinical trial, genomic, and imaging data.

### 2. RSF Meeting Record

| **Meeting date** | **Location** |
|---|---|
| July 23, 2019 | Teleconference (TC) |
| August 28, 2019 | TC |
| September 18, 2019 | TC |
| October 9, 2019 | TC |
| November 25, 2019 | TC |
| December 11, 2019 | TC |
| January 21, 2020 | In person (London, UK) |
| February 27, 2020 | In person (Palm Springs, CA) |
| March 16, 2020 | TC |
| April 27, 2020 | TC |
| May 18, 2020 | TC |
| June 25, 2020 | TC |
| July 13, 2020 | TC |
| September 21, 2020 | TC |
| October 12, 2020 | TC |
| November 9, 2020 | TC |
| December 14, 2020 | TC |
| January 13, 2021 | TC |
| February 22, 2021 | TC |
| March 22, 2021 | TC |
| July 14, 2021 | TC |

### 3. Selection of Landmarks

We conducted a literature review to generate a list of all biomarkers and assessments (collectively, "assessments") used in HD research via a systematic review of the published literature based on work led by Dr. Tiago Mestre. The literature review was conducted by Rancho Biosciences, an independent research organization that performs data curation services, including data search and analysis using algorithmic methods that include natural language processing. The goal was to identify human studies done in Huntington's disease (HD) subjects for biomarker, clinical, and functional decline. In addition to the 140 references listed in Dr. Mestre's 2014 report, Rancho ran two literature searches in PubMed to find relevant articles published since 2014. The following keywords were used:

| Search # | Keyword 1 | Keyword 2 | Keyword 3 | Keyword 4 | Number of publications returned by PubMed | Number of relevant publications | % of relevant publications |
|---|---|---|---|---|---|---|---|
| 1 | human | Huntington disease<br>OR<br>Huntington's disease | participants<br>OR<br>patients | - | 993 | 209 | 21% |
| 2 | human | Huntington disease<br>OR<br>Huntington's disease | participants<br>OR<br>patients | marker<br>OR<br>biomarker | 94 | 59 | 62.8% |

A total of 389 publications were annotated. One record per assessment was created, resulting in 2787 records, since some publications used more than one assessment. The records were divided in the following categories:

Biomarker:

- metabolic biomarkers (blood)
- proteomic (blood)
- biochemical (blood)
- CSF biomarkers
- urine biomarkers
- radiologic biomarkers (MRI/PET/ CT)
- genetic biomarkers (genetic modifiers/ influencing genes)

Clinical and functional decline:

- Changes in clinical status:
  - oculomotor
  - motor
  - psychological
  - cognitive measures
  - electrophysiology
- Changes in functional status (TFC, UHDRS, SF-36, etc.)

This list of candidate assessments was filtered to only include those unique assessments evaluated in studies of high methodologic robustness, defined by a) longitudinal design; and b) large sample size (n > 100). The studies fulfilling these criteria (listed in Table S1) identified a total of 171 unique assessments, presented in Table S2.

***Table S1. Publications with longitudinal design and large sample size (n > 100)***

| |
|---|
| Auinger, P., Kieburtz, K. & McDermott, M. P. The relationship between uric acid levels and Huntington's disease progression. *Mov Disord* **25**, 224-228, doi:10.1002/mds.22907 (2010). |
| Aylward, E. H. *et al.* Longitudinal change in regional brain volumes in prodromal Huntington disease. *J Neurol Neurosurg Psychiatry* **82**, 405-410, doi:10.1136/jnnp.2010.208264 (2011). |
| Baake, V. *et al.* Cognitive decline in Huntington's disease expansion gene carriers. *Cortex* **95**, 51-62, doi:10.1016/j.cortex.2017.07.017 (2017). |
| Boileau, N. R. *et al.* Reliability and Validity of the HD-PRO-TriadTM, a Health-Related Quality of Life Measure Designed to Assess the Symptom Triad of Huntington's Disease. *J Huntingtons Dis* **6**, 201-215, doi:10.3233/JHD-170238 (2017). |
| Borowsky, B. *et al.* 8OHdG is not a biomarker for Huntington disease state or progression. *Neurology* **80**, 1934-1941, doi:10.1212/WNL.0b013e318293e1a1 (2013). |
| D, J. F. *et al.* Multimodal imaging biomarkers in premanifest and early Huntington's disease: 30-month IMAGE-HD data. *Br J Psychiatry* **208**, 571-578, doi:10.1192/bjp.bp.114.156588 (2016). |
| Dominguez, D. J. *et al.* Longitudinal changes in the fronto-striatal network are associated with executive dysfunction and behavioral dysregulation in Huntington's disease: 30 months IMAGE-HD data. *Cortex* **92**, 139-149, doi:10.1016/j.cortex.2017.04.001 (2017). |
| Downing, N. R. *et al.* WHODAS 2.0 in prodromal Huntington disease: measures of functioning in neuropsychiatric disease. *Eur J Hum Genet* **22**, 958-963, doi:10.1038/ejhg.2013.275 (2014). |
| Epping, E. A. *et al.* Longitudinal Psychiatric Symptoms in Prodromal Huntington's Disease: A Decade of Data. *Am J Psychiatry* **173**, 184-192, doi:10.1176/appi.ajp.2015.14121551 (2016). |
| Gregory, S. *et al.* Testing a longitudinal compensation model in premanifest Huntington's disease. *Brain* **141**, 2156-2166, doi:10.1093/brain/awy122 (2018). |
| Hobbs, N. Z. *et al.* Short-interval observational data to inform clinical trial design in Huntington's disease. *J Neurol Neurosurg Psychiatry* **86**, 1291-1298, doi:10.1136/jnnp-2014-309768 (2015). |
| Jacobs, M. *et al.* Progression of motor subtypes in Huntington's disease: a 6-year follow-up study. *J Neurol* **263**, 2080-2085, doi:10.1007/s00415-016-8233-x (2016). |

Labuschagne, I. *et al.* Visuospatial Processing Deficits Linked to Posterior Brain Regions in Premanifest and Early Stage Huntington's Disease. *J Int Neuropsychol Soc* **22**, 595-608, doi:10.1017/S1355617716000321 (2016).

Lambrecq, V. *et al.* Evolution of brain gray matter loss in Huntington's disease: a meta-analysis. *Eur J Neurol* **20**, 315-321, doi:10.1111/j.1468-1331.2012.03854.x (2013).

Li, K., Furr-Stimming, E., Paulsen, J. S., Luo, S. & Group, P.-H. I. o. t. H. S. Dynamic Prediction of Motor Diagnosis in Huntington's Disease Using a Joint Modeling Approach. *J Huntingtons Dis* **6**, 127-137, doi:10.3233/JHD-170236 (2017).

Long, J. D. *et al.* Tracking motor impairments in the progression of Huntington's disease. *Mov Disord* **29**, 311-319, doi:10.1002/mds.25657 (2014).

Meyer, C. *et al.* Rate of change in early Huntington's disease: a clinicometric analysis. *Mov Disord* **27**, 118-124, doi:10.1002/mds.23847 (2012).

Moss, D. J. H. *et al.* Identification of genetic variants associated with Huntington's disease progression: a genome-wide association study. *Lancet Neurol* **16**, 701-711, doi:10.1016/S1474-4422(17)30161-8 (2017).

Nopoulos, P. C. *et al.* Cerebral cortex structure in prodromal Huntington disease. *Neurobiol Dis* **40**, 544-554, doi:10.1016/j.nbd.2010.07.014 (2010).

Orth, M. *et al.* Natural variation in sensory-motor white matter organization influences manifestations of Huntington's disease. *Hum Brain Mapp* **37**, 4615-4628, doi:10.1002/hbm.23332 (2016).

Paulsen, J. S. *et al.* Detection of Huntington's disease decades before diagnosis: the Predict-HD study. *J Neurol Neurosurg Psychiatry* **79**, 874-880, doi:10.1136/jnnp.2007.128728 (2008).

Paulsen, J. S. *et al.* Striatal and white matter predictors of estimated diagnosis for Huntington disease. *Brain Res Bull* **82**, 201-207, doi:10.1016/j.brainresbull.2010.04.003 (2010).

Paulsen, J. S., Smith, M. M., Long, J. D., investigators, P. H. & Coordinators of the Huntington Study, G. Cognitive decline in prodromal Huntington Disease: implications for clinical trials. *J Neurol Neurosurg Psychiatry* **84**, 1233-1239, doi:10.1136/jnnp-2013-305114 (2013).

Rowe, K. C. *et al.* Self-paced timing detects and tracks change in prodromal Huntington disease. *Neuropsychology* **24**, 435-442, doi:10.1037/a0018905 (2010).

Ruiz-Idiago, J. M. *et al.* Spanish Validation of the Problem Behaviors Assessment-Short (PBA-s) for Huntington's Disease. *J Neuropsychiatry Clin Neurosci* **29**, 31-38, doi:10.1176/appi.neuropsych.16020025 (2017).

Rupp, J. *et al.* Progression in prediagnostic Huntington disease. *J Neurol Neurosurg Psychiatry* **81**, 379-384, doi:10.1136/jnnp.2009.176982 (2010).

Saleh, N. *et al.* High insulinlike growth factor I is associated with cognitive decline in Huntington disease. *Neurology* **75**, 57-63, doi:10.1212/WNL.0b013e3181e62076 (2010).

Salem, L. *et al.* Insulin-Like Growth Factor-1 but Not Insulin Predicts Cognitive Decline in Huntington's Disease. *PLoS One* **11**, e0162890, doi:10.1371/journal.pone.0162890 (2016).

Shaffer, J. J. *et al.* Longitudinal diffusion changes in prodromal and early HD: Evidence of white-matter tract deterioration. *Hum Brain Mapp* **38**, 1460-1477, doi:10.1002/hbm.23465 (2017).

Squitieri, F. *et al.* Distinct brain volume changes correlating with clinical stage, disease progression rate, mutation size, and age at onset prediction as early biomarkers of brain atrophy in Huntington's disease. *CNS Neurosci Ther* **15**, 1-11, doi:10.1111/j.1755-5949.2008.00068.x (2009).

| |
|---|
| Stout, J. C. *et al.* Evaluation of longitudinal 12 and 24 month cognitive outcomes in premanifest and early Huntington's disease. *J Neurol Neurosurg Psychiatry* **83**, 687-694, doi:10.1136/jnnp-2011-301940 (2012). |
| Tabrizi, S. J. *et al.* Potential endpoints for clinical trials in premanifest and early Huntington's disease in the TRACK-HD study: analysis of 24 month observational data. *Lancet Neurol* **11**, 42-53, doi:10.1016/S1474-4422(11)70263-0 (2012). |
| Tabrizi, S. J. *et al.* Biological and clinical changes in premanifest and early stage Huntington's disease in the TRACK-HD study: the 12-month longitudinal analysis. *Lancet Neurol* **10**, 31-42, doi:10.1016/S1474-4422(10)70276-3 (2011). |
| Tabrizi, S. J. *et al.* Predictors of phenotypic progression and disease onset in premanifest and early-stage Huntington's disease in the TRACK-HD study: analysis of 36-month observational data. *Lancet Neurol* **12**, 637-649, doi:10.1016/S1474-4422(13)70088-7 (2013). |
| Thompson, J. C. *et al.* Longitudinal evaluation of neuropsychiatric symptoms in Huntington's disease. *J Neuropsychiatry Clin Neurosci* **24**, 53-60, doi:10.1176/appi.neuropsych.11030057 (2012). |
| van Duijn, E. *et al.* Course of irritability, depression and apathy in Huntington's disease in relation to motor symptoms during a two-year follow-up period. *Neurodegener Dis* **13**, 9-16, doi:10.1159/000343210 (2014). |
| van Vugt, J. P. *et al.* Objective assessment of motor slowness in Huntington's disease: clinical correlates and 2-year follow-up. *Mov Disord* **19**, 285-297, doi:10.1002/mds.10718 (2004). |
| van Vugt, J. P. *et al.* Quantitative assessment of daytime motor activity provides a responsive measure of functional decline in patients with Huntington's disease. *Mov Disord* **16**, 481-488, doi:10.1002/mds.1097 (2001). |
| Williams, J. K. *et al.* Everyday cognition in prodromal Huntington disease. *Neuropsychology* **29**, 255-267, doi:10.1037/neu0000102 (2015). |


***Table S2. Unique assessments with longitudinal design and n>100.***

| Category | Assessment |
|---|---|
| Biochemical (blood) | Plasma GH |
| Cognitive | Benton Facial Recognition |
| Cognitive | California Verbal Learning Test |
| Cognitive | Categorical Fluency |
| Cognitive | Circle Tracing |
| Cognitive | COWAT |
| Cognitive | Cued Movement Sequencing |
| Cognitive | Emotion recognition (negative emotions, number correct) |
| Cognitive | Global Cognitive Composite |
| Cognitive | HD-PRO-TRIAD score Cognition |
| Cognitive | Indirect circle tracing annulus length (log cm) |
| Cognitive | Letter Number Sequencing |
| Cognitive | Map Search task |

| | |
|---|---|
| Cognitive | Matrix Reasoning |
| Cognitive | Mental Rotation task percent correct |
| Cognitive | Odour identification |
| Cognitive | Paced Tapping |
| Cognitive | SDMT |
| Cognitive | Shifting response set task |
| Cognitive | Spot the Change 5 s (number correct, corrected for guessing) |
| Cognitive | Stroop Color |
| Cognitive | The Everyday Cognition scales (ecog) |
| Cognitive | Towers Four Disk Condition |
| Cognitive | Trail Making Test |
| Cognitive | Two-choice Response Time |
| Cognitive | UPSIT smell identification (number correct) |
| Cognitive | Verbal Fluency |
| Cognitive | Verbal Working Memory (d-prime) |
| Cognitive | WAIS-R/WAIS III |
| Cognitive | Word list learning (HVLT) |
| Electrophysiology | SEP N20 latency (ms) |
| Electrophysiology | Sep n20/p25 amplitude (sqr mv) |
| Functional changes | Euroqol5d |
| Functional changes | Quality of life (total score) |
| Functional changes | Rand-12 mental health score |
| Functional changes | Rand-12 physical health score |
| Functional changes | Sf36 (total score) |
| Functional changes | Total functional capacity |
| Functional changes | UHDRS functional checklist |
| Functional changes | UHDRS independence scale |
| Metabolic (blood) | Uric acid |
| Motor | Activity level |
| Motor | Activity level adjusted for periods without movement |
| Motor | Alternating-Thumbs Tapping self-paced timing |
| Motor | Chorea Index |
| Motor | Consistency in self-timed finger tapping |
| Motor | Dominant Hand Index Finger Tapping self-paced timing |
| Motor | Gaitrite stance time normal speed (log mean CV) |
| Motor | Grip Force |

| | |
|---|---|
| Motor | H Scan |
| Motor | HD-PRO-TRIAD score Motor |
| Motor | Motor speed: Movement time (MT; 1n msec) |
| Motor | Motor speed: Reaction time (RT; 1n msec) |
| Motor | Movement index |
| Motor | Neuroqol Lower Extremity Function |
| Motor | Neuroqol ltpper Extremity Function |
| Motor | Position index (log) |
| Motor | Speeded Tapping |
| Motor | Tongue Force Variability |
| Motor | Total Motor Score |
| Oculomotor | Anti-saccade |
| Oculomotor | Memory guided task |
| Oculomotor | Prosaccade latency precision (1/SD in 1/ms) |
| Proteomic (blood) | Plasma igf/insulin/etc. |
| Psychological | Apathy |
| Psychological | Depression |
| Psychological | Frontal Systems Behavior Scale (frsbe) |
| Psychological | HD-PRO-TRIAD score Emotional and Behavioral Dyscontrol |
| Psychological | Irritability |
| Psychological | PBA affect (points on PBA scale) |
| Psychological | PBA anger or aggression (square root of points on PBA scale) |
| Psychological | PBA apathy (points on PBA scale) |
| Psychological | PBA composite behaviour score (square root of points on PBA scale) |
| Psychological | PBA irritability (points on PBA scale) |
| Psychological | Problem Behaviors Assessment: Total Score |
| Psychological | Problem Behaviors Assessment-Short: Angry/aggressive behavior |
| Psychological | Problem Behaviors Assessment-Short: Anxiety |
| Psychological | Problem Behaviors Assessment-Short: Delusions/paranoid thinking |
| Psychological | Problem Behaviors Assessment-Short: Depressed mood |
| Psychological | Problem Behaviors Assessment-Short: Disoriented behaviour |
| Psychological | Problem Behaviors Assessment-Short: Hallucinations |
| Psychological | Problem Behaviors Assessment-Short: obsessive-compulsive behavior |
| Psychological | Problem Behaviors Assessment-Short: Perseverative thinking behavior |
| Psychological | Problem Behaviors Assessment-Short: Suicidal ideation |
| Psychological | PROMIS Anger |

| | |
|---|---|
| Psychological | PROMIS Anxiety |
| Psychological | PROMIS Depression |
| Psychological | Schedule of Obsessions, Compulsions and Pathological Impulses, SCOPI |
| Psychological | Symptom Checklist-90-Revised (SCL-90-R): Anxiety |
| Psychological | Symptom Checklist-90-Revised (SCL-90-R): Depression |
| Psychological | Symptom Checklist-90-Revised (SCL-90-R): Global Severity Index |
| Psychological | Symptom Checklist-90-Revised (SCL-90-R): Hostility |
| Psychological | Symptom Checklist-90-Revised (SCL-90-R): Interpersonal Sensitivity |
| Psychological | Symptom Checklist-90-Revised (SCL-90-R): Obsessive–Compulsive |
| Psychological | Symptom Checklist-90-Revised (SCL-90-R): Paranoid Ideation |
| Psychological | Symptom Checklist-90-Revised (SCL-90-R): Phobic Anxiety |
| Psychological | Symptom Checklist-90-Revised (SCL-90-R): Positive Symptom Distress Index |
| Psychological | Symptom Checklist-90-Revised (SCL-90-R): Positive Symptom Total |
| Psychological | Symptom Checklist-90-Revised (SCL-90-R): Psychoticism |
| Psychological | Symptom Checklist-90-Revised (SCL-90-R): Somatization |
| Psychological | UHDRS Psychiatric Mean (SD) |
| Psychological | UHDRS Total Behavioural Score |
| Psychological | WHODAS scores |
| Radiologic | Axial diffusivity (AD) Cortico-spinal tract |
| Radiologic | Axial diffusivity (AD) Motor cortex – motor thalamus tract |
| Radiologic | Axial diffusivity (AD) Premotor cortex – motor thalamus tract |
| Radiologic | Axial diffusivity (AD) Somatosensory cortex – Sensory thalamus tract |
| Radiologic | Blood-oxygen level dependent (BOLD) signal changes in anterior cingulate cortex |
| Radiologic | Blood-oxygen level dependent (BOLD) signal changes in medial prefrontal cortex |
| Radiologic | Blood-oxygen level dependent (BOLD) signal changes in posterior cingulate/precuneus |
| Radiologic | Blood-oxygen level dependent (BOLD) signal changes in putamen |
| Radiologic | Blood-oxygen level dependent (BOLD) signal changes in right dorsolateral prefrontal cortex |
| Radiologic | Blood-oxygen level dependent (BOLD) signal changes in striatum |
| Radiologic | Caudate atrophy |
| Radiologic | Cerebellum volume |
| Radiologic | Changes in microstructural diffusivity |
| Radiologic | Cortical thickness |
| Radiologic | CSF volume |
| Radiologic | Decreased fractional anisotropy (FA) in pre-motor area and putamen |
| Radiologic | Effective connection from left dorsolateral prefrontal cortex to right dorsolateral prefrontal cortex |
| Radiologic | Effective connection from left premotor cortex to right premotor cortex |

| | |
|---|---|
| Radiologic | Effective connection from right dorsolateral prefrontal cortex to left dorsolateral prefrontal cortex |
| Radiologic | Effective connection from right premotor cortex to left posterior parietal cortex |
| Radiologic | Effective connection from right premotor cortex to left premotor cortex |
| Radiologic | Fractional Anisotropy (FA) Cortico-spinal tract |
| Radiologic | Fractional Anisotropy (FA) Motor cortex – motor thalamus tract |
| Radiologic | Fractional Anisotropy (FA) Premotor cortex – motor thalamus tract |
| Radiologic | Fractional Anisotropy (FA) Somatosensory cortex – Sensory thalamus tract |
| Radiologic | Functional connection between left dorsolateral prefrontal cortex and right lateral occipital cortex |
| Radiologic | Grey matter volume |
| Radiologic | Increased axial diffusivity (AD) in pre-motor area and putamen |
| Radiologic | Increased mean diffusivity (MD) in pre-motor area and putamen |
| Radiologic | Increased radial diffusivity (RD) in pre-motor area and putamen |
| Radiologic | Macrostructural volume |
| Radiologic | Mean cortical volumes of bank Superior Temporal Sulcus region |
| Radiologic | Mean cortical volumes of cuneus region |
| Radiologic | Mean cortical volumes of fusiform region |
| Radiologic | Mean cortical volumes of inferior parietal region |
| Radiologic | Mean cortical volumes of inferior temporal region |
| Radiologic | Mean cortical volumes of lateral occipital region |
| Radiologic | Mean cortical volumes of lingual region |
| Radiologic | Mean cortical volumes of middle temporal region |
| Radiologic | Mean cortical volumes of para-central region |
| Radiologic | Mean cortical volumes of pars opercularis region |
| Radiologic | Mean cortical volumes of post-central region |
| Radiologic | Mean cortical volumes of pre-central region |
| Radiologic | Mean cortical volumes of pre-cuneus region |
| Radiologic | Mean cortical volumes of rostral anterior cingulate region |
| Radiologic | Mean cortical volumes of rostral middle frontal region |
| Radiologic | Mean cortical volumes of superior frontal region |
| Radiologic | Mean cortical volumes of superior parietal region |
| Radiologic | Mean cortical volumes of superior temporal region |
| Radiologic | Mean cortical volumes of supra-marginal region |
| Radiologic | Mean cortical volumes of transverse temporal region |
| Radiologic | Radial diffusivity (RD) Cortico-spinal tract |
| Radiologic | Radial diffusivity (RD) Motor cortex – motor thalamus tract |
| Radiologic | Radial diffusivity (RD) Premotor cortex – motor thalamus tract |

| | |
|---|---|
| Radiologic | Radial diffusivity (RD) Somatosensory cortex – Sensory thalamus tract |
| Radiologic | Surface area of superior temporal cortical region |
| Radiologic | Surface area of cuneus cortical region |
| Radiologic | Surface area of postcentral cortical region |
| Radiologic | Surface area of precentral cortical region |
| Radiologic | Surface area of transverse temporal cortical region |
| Radiologic | Total striatum volume |
| Radiologic | Volume of the putamen |
| Radiologic | Volume of cerebral white matter |
| Radiologic | Volume of CSF |
| Radiologic | Volume of the globus pallidus |
| Radiologic | Volume of the striatum |
| Radiologic | Volume of the thalamus |
| Radiologic | Volume of whole brain |

To ensure that classification in a clinical stage has prognostic value, the landmarks that define the stages must be prognostic. Therefore, out of this smaller subset of 171, we selected the 27 assessments for which there existed evidence of prognostic value for events later in the course of the disease, presented in Table S3. An assessment was considered "strong" if it had prognostic value demonstrated in two or more models or in one model that included at least two datasets.

***Table S3. Assessments with demonstrated prognostic value.***

| **Assessment** | **Strong?** | **Assessment** | **Strong?** | **Assessment** | **Strong?** |
|---|---|---|---|---|---|
| Apathy (PBA) | N | Odor (UPSIT) | N | Tapping (TMS item) | Y |
| Caudate Volume | Y | Pallidus Volume | Y | TFC | Y |
| Chorea (TMS) | Y | Plasma GH | N | Thalamus Volume | Y |
| Emotion Recognition | N | Putamen Volume | Y | TMS | Y |
| Gray Matter Volume | N | SDMT | Y | Tongue Force Variability | N |
| HVLT | N | Speed Tapping | N | Uric Acid | N |
| Independence Scale | Y | Spot the Change | Y | Ventricular Volume | Y |
| NfL Plasma | N | Striatum Volume | N | Verbal Fluency | Y |
| Occupation (TFC) | Y | Stroop Color | Y | White Matter Volume | Y |

Finally, the modeling evidence supporting the prognostic value of the six chosen landmarks is presented in Table S4.

***Table S4. Modeling evidence for the prognostic value of the six chosen landmarks.***

| **Landmark** | **Modeling evidence** |
|---|---|
| Caudate Volume | Wijeratne, P. A. *et al.* An image-based model of brain volume biomarker changes in Huntington's disease. *Ann Clin Transl Neurol* **5**, 570-582, doi:10.1002/acn3.558 (2018);[1]<br><br>Wijeratne, P. A. *et al.* Robust Markers and Sample Sizes for Multicenter Trials of Huntington Disease. *Ann Neurol* **87**, 751-762, doi:10.1002/ana.25709 (2020);[2]<br><br>Sun Z, Li Y, Ghosh S, et al. A Data-Driven Method for Generating Robust Symptom Onset Indicators in Huntington's Disease Registry Data. *Amia Annu Symposium Proc Amia Symposium*. 2017;2017:1635-1644;[3]<br><br>Ghosh S, Sun Z, Li Y, et al. An Exploration of Latent Structure in Observational Huntington's Disease Studies. *AMIA Joint Summits on Translational Science proceedings AMIA Joint Summits on Translational Science*. 2017;2017:92-102;[4]<br><br>Tabrizi, S. J. *et al.* Biological and clinical manifestations of Huntington's disease in the longitudinal TRACK-HD study: cross-sectional analysis of baseline data. *Lancet Neurol* **8**, 791-801, doi:10.1016/S1474-4422(09)70170-X (2009).[5] |
| Putamen Volume | Wijeratne, P. A. *et al.* An image-based model of brain volume biomarker changes in Huntington's disease. *Ann Clin Transl Neurol* **5**, 570-582, doi:10.1002/acn3.558 (2018);[1] |

| | |
|---|---|
| | Wijeratne, P. A. *et al.* Robust Markers and Sample Sizes for Multicenter Trials of Huntington Disease. *Ann Neurol* **87**, 751-762, doi:10.1002/ana.25709 (2020);[2]<br><br>Sun Z, Li Y, Ghosh S, et al. A Data-Driven Method for Generating Robust Symptom Onset Indicators in Huntington's Disease Registry Data. *Amia Annu Symposium Proc Amia Symposium*. 2017;2017:1635-1644;[3]<br><br>Ghosh S, Sun Z, Li Y, et al. An Exploration of Latent Structure in Observational Huntington's Disease Studies. *AMIA Joint Summits on Translational Science proceedings AMIA Joint Summits on Translational Science*. 2017;2017:92-102;[4]<br><br>Tabrizi, S. J. *et al.* Biological and clinical manifestations of Huntington's disease in the longitudinal TRACK-HD study: cross-sectional analysis of baseline data. *Lancet Neurol* **8**, 791-801, doi:10.1016/S1474-4422(09)70170-X (2009);[5]<br><br>Paulsen, J. S. *et al.* Prediction of manifest Huntington's disease with clinical and imaging measures: a prospective observational study. *Lancet Neurol* **13**, 1193-1201, doi:10.1016/S1474-4422(14)70238-8 (2014).[6] |
| TMS | Wijeratne, P. A. *et al.* An image-based model of brain volume biomarker changes in Huntington's disease. *Ann Clin Transl Neurol* **5**, 570-582, doi:10.1002/acn3.558 (2018);[1]<br><br>Wijeratne, P. A. *et al.* Robust Markers and Sample Sizes for Multicenter Trials of Huntington Disease. *Ann Neurol* **87**, 751-762, doi:10.1002/ana.25709 (2020);[2]<br><br>Tabrizi, S. J. *et al.* Predictors of phenotypic progression and disease onset in premanifest and early-stage Huntington's disease in the TRACK-HD study: analysis of 36-month observational data. *Lancet Neurol* **12**, 637-649, doi:10.1016/S1474-4422(13)70088-7 (2013);[7]<br><br>Long, J. D., Paulsen, J. S., Investigators, P.-H. & Coordinators of the Huntington Study, G. Multivariate prediction of motor diagnosis in Huntington's disease: 12 years of PREDICT-HD. *Mov Disord* **30**, 1664-1672, doi:10.1002/mds.26364 (2015);[8]<br><br>Li, F. *et al.* Predicting the Risk of Huntington's Disease with Multiple Longitudinal Biomarkers. *J Huntingtons Dis* **8**, 323-332, doi:10.3233/JHD-190345 (2019);[9]<br><br>Paulsen, J. S. *et al.* Challenges assessing clinical endpoints in early Huntington disease. *Mov Disord* **25**, 2595-2603, doi:10.1002/mds.23337 (2010).[10] |
| SDMT | Tabrizi, S. J. *et al.* Predictors of phenotypic progression and disease onset in premanifest and early-stage Huntington's disease in the TRACK-HD study: analysis of 36-month observational data. *Lancet Neurol* **12**, 637-649, doi:10.1016/S1474-4422(13)70088-7 (2013);[7]<br><br>Long, J. D., Paulsen, J. S., Investigators, P.-H. & Coordinators of the Huntington Study, G. Multivariate prediction of motor diagnosis in Huntington's disease: 12 years of PREDICT-HD. *Mov Disord* **30**, 1664-1672, doi:10.1002/mds.26364 (2015);[8]<br><br>Li, F. *et al.* Predicting the Risk of Huntington's Disease with Multiple Longitudinal Biomarkers. *J Huntingtons Dis* **8**, 323-332, doi:10.3233/JHD-190345 (2019);[9]<br><br>Paulsen, J. S. *et al.* Challenges assessing clinical endpoints in early Huntington disease. *Mov Disord* **25**, 2595-2603, doi:10.1002/mds.23337 (2010).[10] |
| TFC | Li, F. *et al.* Predicting the Risk of Huntington's Disease with Multiple Longitudinal Biomarkers. *J Huntingtons Dis* **8**, 323-332, doi:10.3233/JHD-190345 (2019);[9] |

| | |
|---|---|
| | Beglinger, L. J. *et al.* Clinical predictors of driving status in Huntington's disease. *Mov Disord* **27**, 1146-1152, doi:10.1002/mds.25101 (2012).[11] |
| Independence Scale | Sun Z, Li Y, Ghosh S, et al. A Data-Driven Method for Generating Robust Symptom Onset Indicators in Huntington's Disease Registry Data. *Amia Annu Symposium Proc Amia Symposium*. 2017;2017:1635-1644;[3]<br><br>Ghosh S, Sun Z, Li Y, et al. An Exploration of Latent Structure in Observational Huntington's Disease Studies. *AMIA Joint Summits on Translational Science proceedings AMIA Joint Summits on Translational Science*. 2017;2017:92-102.[4] |

### 4. Statistical Methods

#### A. Reduced Penetrance

Penetrance is defined as the proportion of individuals with a given genotype who exhibit the phenotype associated with that genotype.[12] For this analysis, the genotype was CAG length as measured in the blood, and the phenotype was HD clinical motor diagnosis, defined as the highest score on the UHDRS diagnostic confidence level (DCL), or DCL = 4 (DCL4).

We considered complete penetrance to exist for a given CAG length when all individuals of that length reach DCL4 in a reasonable expected lifespan. Reduced penetrance of a CAG length means that not every individual of that CAG length has reached DCL = 4 in a reasonable expected lifespan. Therefore, interest was focused on individuals who were older than the expected lifespan without DCL4.

There were two strategies for the reduced penetrance analysis; the first was descriptive and the second was inferential (based on statistical modeling). The first approach was to examine the proportion of DCL4 by CAG. CAG lengths were compared for all ages and considered in relation to the reference point of 82 years of age. This reference is the approximate United Nations life expectancy estimate of a western European person (combined males and females) for 2020-2025.[13] This analysis focused on the most recent visit of each individual in Enroll-HD,[14] PREDICT-HD,[8,15-17] and TRACK-HD.[12,27,28] The counts and age statistics by CAG for the combined sample are shown in Table S5.

***Table S5. Counts of participants and age range by CAG expansion at most recent study visit.***

| **CAG** | **N** | **Minimum Age** | **Maximum Age** |
|---|---|---|---|
| 36 | 40 | 23.03 | 78.99 |
| 37 | 51 | 27.45 | 80.83 |
| 38 | 165 | 24.30 | 90.42 |
| 39 | 422 | 18.84 | 92.44 |
| 40 | 1216 | 18.65 | 88.88 |
| 41 | 1798 | 18.82 | 85.41 |
| 42 | 2184 | 18.07 | 90.61 |
| 43 | 1869 | 18.16 | 78.72 |

##### i. Descriptive Statistics

Figure 1A in the main text shows the descriptive results.

##### ii. Inferential: Logistic Regression for Longitudinal Data

The second strategy was to fit statistical models to estimate the probability of conversion to DCL4 by CAG. Specifically, we fit longitudinal logistic regression models and estimated the population-average probability of conversion over time. The model was a generalized linear mixed model with logit link and a random intercept. Parameters were estimated using maximum likelihood methods and population-averaged coefficients were obtained using the approximation of Hedeker et al.[18] with the GLMMadaptive package[19] for the R software system. The main question was: for each CAG, at what age range was complete penetrance likely (probability of conversion = 1). In order for the data to have a strong influence on the estimated probabilities, we fit a separate model for each CAG.

Figure 1B in the main text shows the predicted curves of probability of DCL4 by age by CAG. Model-based curves in logistic regression converge to but never actually reach 1. As a proxy for complete penetrance, we considered probability = 0.99 and computed the age at which this probability was estimated to occur. Table S6 shows CAG in the first column and the estimated age at probability = 0.99 in the second column (with the ages being estimated based on the fitted models). The estimated ages for CAG 38 and 39 are greater than 100, whereas the age for CAG 40 is approximately 86. Longer CAG lengths reach probability = 0.99 at estimated ages less than 78.

***Table S6. Estimated age at probability = 0.99 by CAG***

| **CAG** | **Age at Probability = 0.99** |
|---|---|
| 38 | 142.0 |
| 39 | 104.2 |
| 40 | 85.9 |
| 41 | 77.8 |
| 42 | 68.0 |
| 43 | 63.6 |

### B. Participants for Staging System Development and Validation

Four publicly available data sets were used for the main analysis, Enroll-HD (4th periodic data set from 2018),[14] IMAGE-HD (data collected 2008-2013),[20] PREDICT-HD (data collected 2002-2014),[6] and TRACK-HD/ON (data collected 2008-2014)[7,21]. Table S7 shows the key variables and sample sizes for controls (CAG < 36) and cases (CAG ≥ 40).

The controls were from all four studies, most being family members of cases (there was a small number of unrelated community controls). The controls totaled $N = 4636$ with 13300 visits. The majority had two or more visits (72.7%) with most individuals having between 2 and 7 visits (maximum = 15).

To focus on transition through all the stages, the cases were only included from the three studies that had imaging data: IMAGE-HD, PREDICT-HD, TRACK-HD/ON. The number of participants and observations was sparse for CAG > 50 and therefore the range of 40-50 CAG was used for the analysis. The cases totaled $N = 1107$ with 3176 visits. The majority had two or more visits (75.5%) with most individuals having between 2 and 7 visits, with a maximum of 9. The controls outnumbered the cases; the former were used to establish cut-offs on each variable in isolation rather than to study progression through the Stages. Table S7 shows the baseline characteristics of the participants.

***Table S7. Baseline characteristics of participants.***

| Status | Study | Var | N | Mean | SD | Min | Max | Q1 | Q2 | Q3 |
|---|---|---|---|---|---|---|---|---|---|---|
| Control | ENROLL | CAG | 3705 | 20.14 | 3.54 | 12.00 | 35.00 | 17.00 | 19.00 | 22.00 |
| Control | IMAGE | CAG | 0 | - | - | - | - | - | - | - |
| Control | PREDICT | CAG | 584 | 20.40 | 3.70 | 12.00 | 35.00 | 18.00 | 19.00 | 22.00 |
| Control | TRACK | CAG | 0 | - | - | - | - | - | - | - |
| Control | ENROLL | >HS | 3699 | 0.60 | 0.49 | 0.00 | 1.00 | 0.00 | 1.00 | 1.00 |
| Control | IMAGE | >HS | 0 | - | - | - | - | - | - | - |
| Control | PREDICT | >HS | 576 | 0.80 | 0.40 | 0.00 | 1.00 | 1.00 | 1.00 | 1.00 |
| Control | TRACK | >HS | 311 | 0.70 | 0.46 | 0.00 | 1.00 | 0.00 | 1.00 | 1.00 |
| Control | ENROLL | Female | 3705 | 0.61 | 0.49 | 0.00 | 1.00 | 0.00 | 1.00 | 1.00 |
| Control | IMAGE | Female | 36 | 0.67 | 0.48 | 0.00 | 1.00 | 0.00 | 1.00 | 1.00 |
| Control | PREDICT | Female | 584 | 0.65 | 0.48 | 0.00 | 1.00 | 0.00 | 1.00 | 1.00 |
| Control | TRACK | Female | 311 | 0.56 | 0.50 | 0.00 | 1.00 | 0.00 | 1.00 | 1.00 |
| Control | ENROLL | Putamen | 0 | - | - | - | - | - | - | - |
| Control | IMAGE | Putamen | 36 | 6.76 | 0.65 | 5.52 | 8.23 | 6.20 | 6.70 | 7.23 |
| Control | PREDICT | Putamen | 259 | 6.47 | 0.76 | 4.69 | 9.69 | 5.91 | 6.47 | 7.00 |
| Control | TRACK | Putamen | 127 | 7.07 | 1.07 | 5.51 | 11.15 | 6.33 | 6.95 | 7.55 |
| Control | ENROLL | Caudate | 0 | - | - | - | - | - | - | - |
| Control | IMAGE | Caudate | 36 | 4.98 | 0.58 | 3.78 | 6.02 | 4.51 | 4.94 | 5.38 |
| Control | PREDICT | Caudate | 259 | 4.71 | 0.60 | 3.21 | 7.15 | 4.31 | 4.66 | 5.05 |
| Control | TRACK | Caudate | 127 | 5.06 | 0.73 | 3.27 | 7.62 | 4.64 | 4.91 | 5.39 |
| Control | ENROLL | TMS | 3687 | 1.72 | 3.48 | 0.00 | 69.00 | 0.00 | 0.00 | 2.00 |
| Control | IMAGE | TMS | 0 | - | - | - | - | - | - | - |
| Control | PREDICT | TMS | 562 | 2.88 | 3.56 | 0.00 | 25.00 | 0.00 | 2.00 | 4.00 |
| Control | TRACK | TMS | 311 | 1.43 | 1.64 | 0.00 | 7.00 | 0.00 | 1.00 | 2.00 |
| Control | ENROLL | SDMT | 3634 | 49.73 | 12.24 | 0.00 | 97.00 | 43.00 | 51.00 | 58.00 |
| Control | IMAGE | SDMT | 36 | 56.28 | 10.08 | 31.00 | 80.00 | 50.75 | 56.00 | 63.00 |
| Control | PREDICT | SDMT | 556 | 53.91 | 9.49 | 26.00 | 84.00 | 49.00 | 54.00 | 60.00 |
| Control | TRACK | SDMT | 311 | 52.53 | 9.46 | 30.00 | 78.00 | 47.00 | 54.00 | 59.00 |
| Control | ENROLL | TFC | 3689 | 12.86 | 0.71 | 0.00 | 13.00 | 13.00 | 13.00 | 13.00 |
| Control | IMAGE | TFC | 0 | - | - | - | - | - | - | - |
| Control | PREDICT | TFC | 492 | 12.97 | 0.27 | 8.00 | 13.00 | 13.00 | 13.00 | 13.00 |
| Control | TRACK | TFC | 311 | 12.99 | 0.11 | 12.00 | 13.00 | 13.00 | 13.00 | 13.00 |
| Control | ENROLL | IS | 3690 | 99.57 | 2.72 | 45.00 | 100.00 | 100.00 | 100.00 | 100.00 |
| Control | IMAGE | IS | 0 | - | - | - | - | - | - | - |
| Control | PREDICT | IS | 422 | 99.95 | 0.49 | 95.00 | 100.00 | 100.00 | 100.00 | 100.00 |
| Control | TRACK | IS | 311 | 99.94 | 0.56 | 95.00 | 100.00 | 100.00 | 100.00 | 100.00 |

| Status | Study | Var | N | Mean | SD | Min | Max | Q1 | Q2 | Q3 |
|---|---|---|---|---|---|---|---|---|---|---|
| Control | ENROLL | DCL | 3699 | 0.22 | 0.50 | 0.00 | 4.00 | 0.00 | 0.00 | 0.00 |
| Control | IMAGE | DCL | 0 | - | - | - | - | - | - | - |
| Control | PREDICT | DCL | 562 | 0.48 | 0.61 | 0.00 | 3.00 | 0.00 | 0.00 | 1.00 |
| Control | TRACK | DCL | 311 | 0.20 | 0.40 | 0.00 | 1.00 | 0.00 | 0.00 | 0.00 |
| Case | IMAGE | CAG | 67 | 42.66 | 1.89 | 40.00 | 47.00 | 41.00 | 42.00 | 44.00 |
| Case | PREDICT | CAG | 822 | 42.59 | 1.99 | 40.00 | 48.00 | 41.00 | 42.00 | 44.00 |
| Case | TRACK | CAG | 218 | 43.11 | 1.89 | 40.00 | 48.00 | 42.00 | 43.00 | 44.00 |
| Case | IMAGE | >HS | 0 | - | - | - | - | - | - | - |
| Case | PREDICT | >HS | 797 | 0.74 | 0.44 | 0.00 | 1.00 | 0.00 | 1.00 | 1.00 |
| Case | TRACK | >HS | 218 | 0.55 | 0.50 | 0.00 | 1.00 | 0.00 | 1.00 | 1.00 |
| Case | IMAGE | Female | 67 | 0.51 | 0.50 | 0.00 | 1.00 | 0.00 | 1.00 | 1.00 |
| Case | PREDICT | Female | 822 | 0.64 | 0.48 | 0.00 | 1.00 | 0.00 | 1.00 | 1.00 |
| Case | TRACK | Female | 218 | 0.56 | 0.50 | 0.00 | 1.00 | 0.00 | 1.00 | 1.00 |
| Case | IMAGE | Putamen | 67 | 5.05 | 0.98 | 3.12 | 7.07 | 4.25 | 5.01 | 5.77 |
| Case | PREDICT | Putamen | 822 | 5.55 | 1.12 | 2.42 | 10.19 | 4.82 | 5.53 | 6.22 |
| Case | TRACK | Putamen | 218 | 5.25 | 1.30 | 2.97 | 8.85 | 4.29 | 5.12 | 5.98 |
| Case | IMAGE | Caudate | 67 | 3.75 | 0.81 | 2.05 | 5.95 | 3.17 | 3.55 | 4.42 |
| Case | PREDICT | Caudate | 822 | 4.14 | 0.81 | 1.40 | 7.50 | 3.60 | 4.14 | 4.63 |
| Case | TRACK | Caudate | 218 | 3.87 | 0.98 | 1.72 | 6.77 | 3.18 | 3.84 | 4.48 |
| Case | IMAGE | TMS | 67 | 10.10 | 12.97 | 0.00 | 60.00 | 0.00 | 4.00 | 15.00 |
| Case | PREDICT | TMS | 751 | 5.32 | 6.00 | 0.00 | 40.00 | 1.00 | 3.00 | 8.00 |
| Case | TRACK | TMS | 218 | 11.50 | 12.80 | 0.00 | 52.00 | 2.00 | 4.00 | 18.00 |
| Case | IMAGE | SDMT | 67 | 44.04 | 13.09 | 18.00 | 74.00 | 34.50 | 46.00 | 52.00 |
| Case | PREDICT | SDMT | 747 | 50.51 | 11.54 | 16.00 | 92.00 | 44.00 | 50.00 | 58.00 |
| Case | TRACK | SDMT | 218 | 43.87 | 13.52 | 12.00 | 80.00 | 35.00 | 44.00 | 53.00 |
| Case | IMAGE | TFC | 0 | - | - | - | - | - | - | - |
| Case | PREDICT | TFC | 580 | 12.79 | 0.74 | 7.00 | 13.00 | 13.00 | 13.00 | 13.00 |
| Case | TRACK | TFC | 218 | 11.99 | 1.74 | 7.00 | 13.00 | 12.00 | 13.00 | 13.00 |
| Case | IMAGE | IS | 0 | - | - | - | - | - | - | - |
| Case | PREDICT | IS | 580 | 99.46 | 2.55 | 80.00 | 100.00 | 100.00 | 100.00 | 100.00 |
| Case | TRACK | IS | 218 | 96.49 | 7.17 | 70.00 | 100.00 | 95.00 | 100.00 | 100.00 |
| Case | IMAGE | DCL | 65 | 1.45 | 1.67 | 0.00 | 4.00 | 0.00 | 1.00 | 3.00 |
| Case | PREDICT | DCL | 752 | 0.94 | 0.94 | 0.00 | 4.00 | 0.00 | 1.00 | 1.00 |
| Case | TRACK | DCL | 218 | 2.07 | 1.69 | 0.00 | 4.00 | 1.00 | 1.00 | 4.00 |

### C. Cut-off Values

Cut-offs for each variable (Stages 1 to 3) were the extreme quantiles of the variable distribution over time, which were estimated using quantile regression with the repeated measures of only the controls. A random intercept was included to account for the correlation due to repeated measures.

#### i. Quantile Regression for Longitudinal Data

Each variable was treated as the outcome in a separate model. Let $y_{ij}$ be the score of a variable for the $i^{th}$ participant ($i = 1, \ldots, N$) at the $j^{th}$ time point ($j = 1, \ldots, n_i$). Then the model is

$$Q_{y_{ij}}(\tau) = a_i + \alpha^{(\tau)} + \beta_1^{(\tau)} f_1(age_{ij}) + \beta_2^{(\tau)} f_2(age_{ij}) + \beta_3^{(\tau)} f_3(age_{ij}) \qquad \text{(A1)}$$

where $Q_{y_{ij}}(\tau)$ is the quantile function (inverse cumulative function) of $y_{ij}$ given the predictors and the random intercepts ($a_i$), $\alpha^{(\tau)}$ is the fixed intercept, and $\beta_k^{(\tau)}$ is a fixed effect evaluated at quantile $\tau$. Because the functional form of the longitudinal trend is unknown, a natural cubic spline was used with two equally-spaced knots, so that $f_1(\cdot)$, $f_2(\cdot)$, and $f_3(\cdot)$ are the three basis functions.

For SDMT, a binary indicator of education level (0 = high school education or less, 1 = more than high school education) was included to account for initial differences in cognition.[22] For putamen and caudate change, the simpler linear predictor of $a_i + \alpha^{(\tau)} + \beta_1^{(\tau)} t_{ij}$ was used as these volumes are known to have a constant rate of change for controls and cases over the age windows considered here.[6]

A penalization approach to estimation was used that involves $\ell_1$ regularization of the random intercepts with a tuning parameter ($\lambda$) to control the amount of shrinkage.[23] The R package rqpd[24] was used for the analysis.

Figure S1 shows an illustration of the cut-off and scoring process. The observed data for the controls (red) and cases with CAG = 42 (blue) are shown with a thin line connecting repeated visits where applicable. CAG = 42 is selected because this is the length with the largest sample size. The fitted quantile curves (black) are shown for the variables, with SDMT paneled by education (high school education level or less versus more than high school education).

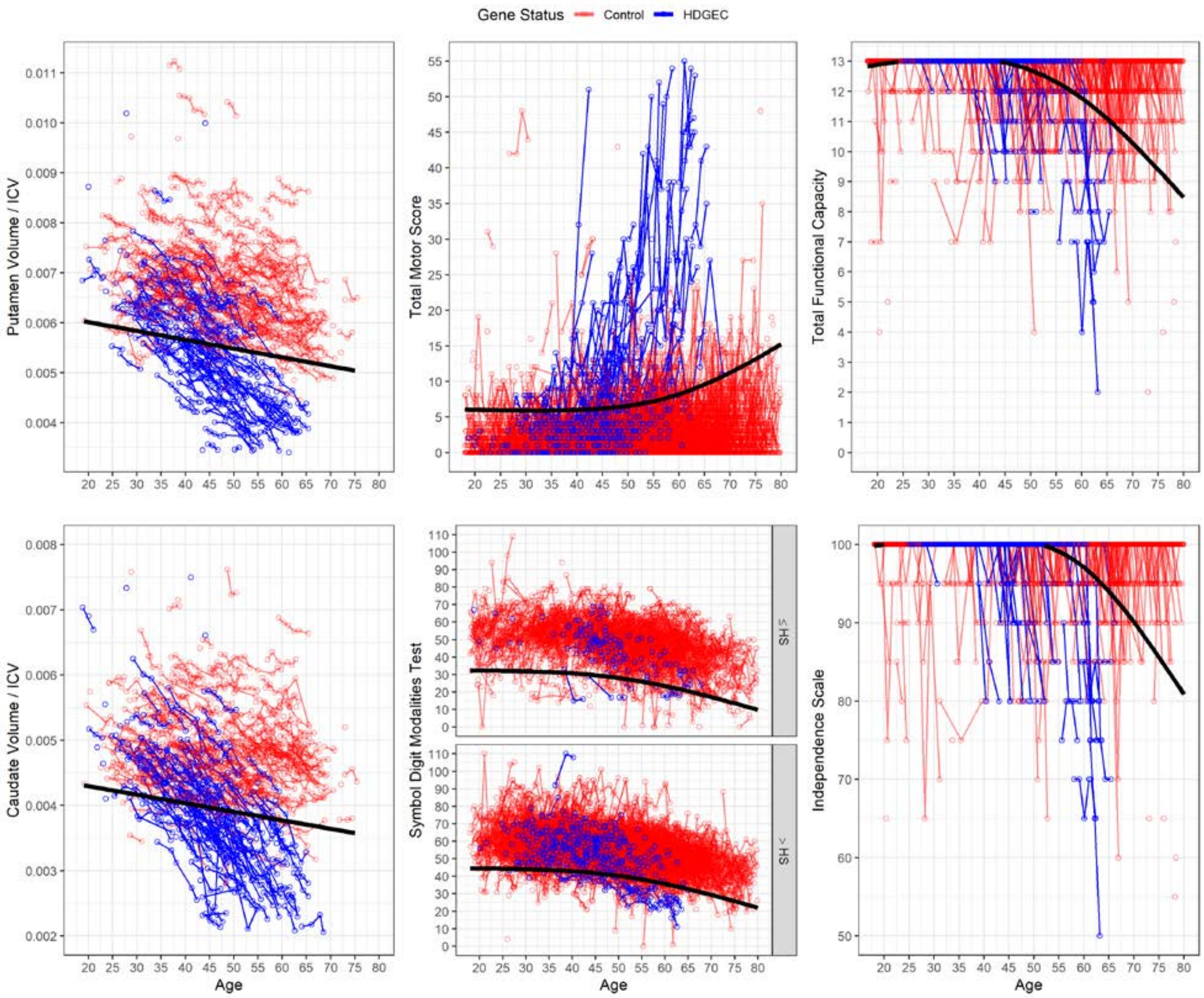


***Figure S1. Observed data of controls (red) and cases with CAG = 42 (blue) of variables for Stages 1, 2, and 3.*** *Solid black lines are the quantile curves based on the controls.*

### D. Scoring and Stage Assignment

Cut-offs were predicted using the estimated fixed effects in Equation A1 (or the alternative models for SDMT and brain volume). The Equation A1 model conditions on the random intercepts. However, our goal was to set quantile cut-offs based on group-level control curves, also known as population-averaged curves. As an alternative to averaging or integrating over the random intercepts, we used a relatively large tuning parameter of $\lambda = 20$ to shrink the random effects towards 0. A similar approach has been used in the estimation of early growth norms.[25] We treated the resulting fixed effects estimates as population-averaged coefficients.

There were six sets of fixed effects for the variables of Stages 1 to 3. We denote the predicted quantile for putamen volume at an age for a participant as $\hat{\mu}^{(.05)}_{\text{put}_{ij}}$, and similarly for the other variables. Explicitly, the predicted cut-off value for putamen volume is denoted as $\hat{\mu}^{(.05)}_{\text{put}_{ij}} = \hat{\alpha}^{(.05)} + \hat{\beta}^{(.05)}_1 \text{age}_{ij}$, where $\text{age}_{ij}$ is the observed visit age of a case. The observed putamen volume (put$_{ij}$) exceeds the cut-off when $\text{put}_{ij} < \hat{\mu}^{(.05)}_{\text{put}_{ij}}$. The same applies for the other variables except TMS, whose observed value exceeds the cut-off when $\text{tms}_{ij} > \hat{\mu}^{(.95)}_{\text{tms}_{ij}}$.

The HD-ISS is cumulative in the sense that a later stage has all the conditions met of the earlier stages. The monotonicity suggests that a stage can be scored based on a sum of binary indicators of Stages 1-3. Suppose the $h^{th}$ condition for a participant is denoted as $c_{hij}, h = 1,2,3,$ and $I\{\cdot\}$ is the indicator function. Then each condition is scored as,

$$
\begin{aligned}
c_{1ij} &= I\{\text{put}_{ij} < \hat{\mu}^{(.05)}_{\text{put}_{ij}} \cup \text{caud}_{ij} < \hat{\mu}^{(.05)}_{\text{caud}_{ij}}\},\\
c_{2ij} &= I\{\text{tms}_{ij} > \hat{\mu}^{(.95)}_{\text{tms}_{ij}} \cup \text{sdmt}_{ij} < \hat{\mu}^{(.05)}_{\text{sdmt}_{ij}}\},\\
c_{3ij} &= I\{\text{tfc}_{ij} < \hat{\mu}^{(.05)}_{\text{tfc}_{ij}} \cup \text{is}_{ij} < \hat{\mu}^{(.05)}_{\text{is}_{ij}}\},
\end{aligned}
$$

where union (∪) indicates that if either relation or both are met in the curly brackets then the indicator takes the value of 1, and 0 otherwise. The observation for a person at a particular point in time is assigned to Stage $s_{ij}$ by summing the condition indicators,

$$
s_{ij} = \sum_{h=1} c_{hij} = c_{1ij} + c_{2ij} + c_{3ij}, \qquad (2)
$$

with $s_{ij} = 0$ for Stage 0. Because of the verified genetic testing, all cases were assumed to be at least Stage 0 in the analysis.

### E. Tail Area Justification

The quantile of $\tau = .95$ was specified for TMS and $\tau = .05$ was used for all other variables. Thus, the tail area of a cut-off was 5% (either upper or lower tail). The 5% tail area is consistent with similar reference-based approaches[26-28] and has been the focus of previous HD research[22]. The statistical justification for a 5% tail area was established by comparing the probability curves for different tail-area models with the probability curve for DCL = 2 (DCL2). DCL2 is the first level at which the rater can attribute motor signs to HD, which is similar to what the TMS indicator is designed for. Therefore, whichever quantile cut-off provides the closest curve fit to DCL2 will be preferred.

Agreement of the probability curves was be assessed using a multivariate generalized linear mixed model with logit link functions and correlated random effects. The TMS model was based on the log odds of $I\{\text{tms}_{ij} > \hat{\mu}^{(\tau)}_{\text{tms}_{ij}}\}$, whereas the DCL model was based on the log odds of $I\{dcl \geq 2\}$. Each linear predictor had a random and fixed intercept, main effects of age and CAG, and the age by CAG interaction. All parameters were estimated simultaneously, and the correlated random effects provided a basis for the variance-covariance matrix among the fixed effects.

Note that the comparison of predicted probability curves is equivalent to the comparison of parameter estimates. To assess the overall similarity of the parameter estimates we considered the linear contrast in which every TMS estimate was compared with its corresponding DCL2 estimate. Then a Wald-type statistic was computed $W = q^T \hat{V} q$, where $q$ is the $8 \times 1$ contrast-coded vector of stacked fixed effects parameter estimates, and $\hat{V}$ is the estimated variance-covariance matrix among all the fixed effects.

Based on research in HD and other diseases, we considered cut-offs for $\tau = .90, .95, .975$. Recall that $\hat{\mu}^{(\tau)}_{\text{tms}_{ij}}$ is based on analysis with controls, whereas here we consider analysis only with cases. For each set of cut-offs, a multivariate model with DCL2 was estimated and the Wald statistics was computed.

Results show $W_{.90} = 78.52$, $W_{.95} = 35.48$, $W_{.975} = 56.20$. Therefore, the cut-off based on $\tau = .95$ provided the closest agreement with DCL2. Figure S2 shows the predicted probability curves of the subject-specific models for CAG = 42. The TMS curve based on the 5% cut-off ($\tau = .95$) shows the closest overall correspondence with the DCL2 curve.

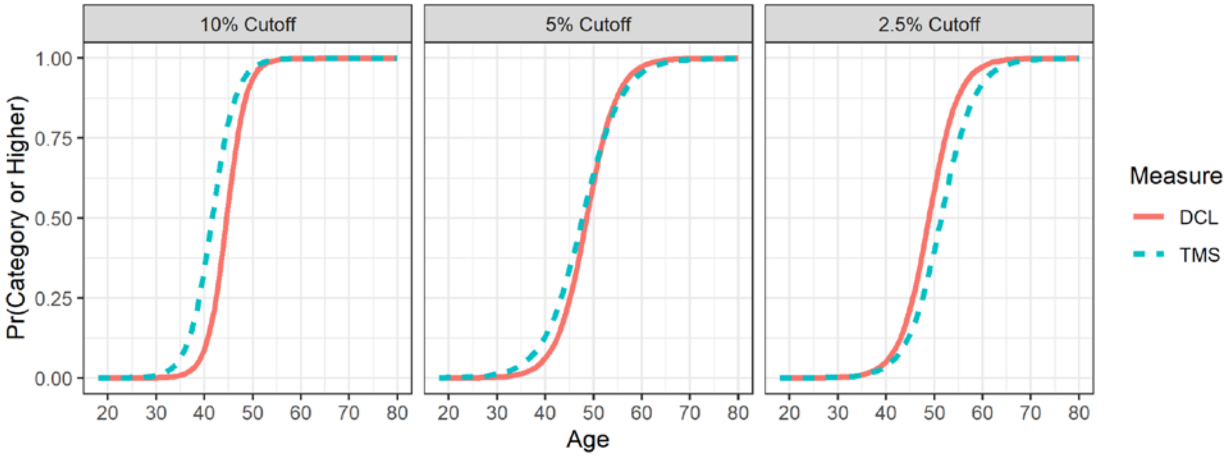


***Figure S2. Predicted Probability curves of subject-specific models for CAG=42.***

### F. Missing Data

As shown in Table S7, several non-imaging variables had missing values, especially for Stage 3. Missing data was handled using multiple imputation under the assumption of an ignorable missing data mechanism.[29] To impute plausible values that were integer and in the permissible range, predictive mean matching was used.[30] Each incomplete variable in Table S7 was imputed based on all the other variables and age using random forest (RF),[31] which accounts for different variable scales, non-linear effects, and high correlations among the predictors. For a missing datum, a pool of potential "donors" is identified whose observed value is close to the predicted value based on the RF. Then one of the $l$ donor values is randomly selected as the imputed value. Based on results of simulation studies, the number of potential donors was set to $l = 10$,[32] and the number of imputed data sets was set to $M = 20$[33]. The frequency distribution was estimated for each imputed data set, and then combining rules[29] were used to pool the results. The R package missRanger was used for the imputation.[34]

A complication was that standard errors (SEs) of the population-averaged coefficients and their estimated probabilities required estimation using the bootstrap (see below). We adopted the approach of bootstrapping each imputed data set to estimate the SEs (and the other parameters), and then pooling the estimates among the imputed data sets using combining rules (the "MI Boot" approach of Schomaker and Heumann 2018[33]). There is evidence that the MI Boot approach provides proper CI coverage when the number of imputed data sets is adequate, $M \geq 20$.[33] The number of bootstrap samples was set to 100, as this appears adequate for estimating SEs in the longitudinal regression models considered below.[19] The bootstrap used here was non-parametric with the entire repeated measures record of a participant being selected when re-sampling in the longitudinal context.

***Table S8. Operational Cut-offs by Age.***

| Age | Condition 1 | | Condition 2 | | | Condition 3 | |
|---|---|---|---|---|---|---|---|
| | Putamen | Caudate | TMS | SDMT[a] | SDMT[b] | TFC | IS |
| 18 | 6.05 | 4.32 | 6 | 32 | 45 | 13 | 100 |
| 19 | 6.03 | 4.31 | 6 | 32 | 45 | 13 | 100 |
| 20 | 6.01 | 4.30 | 6 | 32 | 45 | 13 | 100 |
| 21 | 5.99 | 4.28 | 6 | 32 | 45 | 13 | 100 |
| 22 | 5.98 | 4.27 | 6 | 32 | 45 | 13 | 100 |
| 23 | 5.96 | 4.26 | 6 | 32 | 44 | 13 | 100 |
| 24 | 5.94 | 4.24 | 6 | 32 | 44 | 13 | 100 |
| 25 | 5.92 | 4.23 | 6 | 32 | 44 | 13 | 100 |
| 26 | 5.91 | 4.22 | 6 | 32 | 44 | 13 | 100 |
| 27 | 5.89 | 4.21 | 6 | 32 | 44 | 13 | 100 |
| 28 | 5.87 | 4.19 | 6 | 32 | 44 | 13 | 100 |
| 29 | 5.85 | 4.18 | 6 | 32 | 44 | 13 | 100 |
| 30 | 5.84 | 4.17 | 6 | 32 | 44 | 13 | 100 |
| 31 | 5.82 | 4.15 | 6 | 32 | 44 | 13 | 100 |
| 32 | 5.80 | 4.14 | 6 | 32 | 44 | 13 | 100 |
| 33 | 5.78 | 4.13 | 6 | 32 | 44 | 13 | 100 |
| 34 | 5.76 | 4.11 | 6 | 32 | 44 | 13 | 100 |
| 35 | 5.75 | 4.10 | 6 | 32 | 44 | 13 | 100 |
| 36 | 5.73 | 4.09 | 6 | 32 | 44 | 13 | 100 |
| 37 | 5.71 | 4.07 | 6 | 32 | 44 | 13 | 100 |
| 38 | 5.69 | 4.06 | 6 | 31 | 44 | 13 | 100 |
| 39 | 5.68 | 4.05 | 6 | 31 | 43 | 13 | 100 |
| 40 | 5.66 | 4.04 | 6 | 31 | 43 | 13 | 100 |
| 41 | 5.64 | 4.02 | 6 | 31 | 43 | 13 | 100 |
| 42 | 5.62 | 4.01 | 6 | 31 | 43 | 13 | 100 |
| 43 | 5.61 | 4.00 | 6 | 31 | 43 | 13 | 100 |
| 44 | 5.59 | 3.98 | 6 | 30 | 43 | 13 | 100 |
| 45 | 5.57 | 3.97 | 6 | 30 | 42 | 13 | 100 |
| 46 | 5.55 | 3.96 | 6 | 30 | 42 | 13 | 100 |
| 47 | 5.54 | 3.94 | 6 | 30 | 42 | 13 | 100 |
| 48 | 5.52 | 3.93 | 6 | 29 | 42 | 13 | 100 |
| 49 | 5.50 | 3.92 | 6 | 29 | 41 | 13 | 100 |
| 50 | 5.48 | 3.90 | 6 | 29 | 41 | 13 | 100 |
| 51 | 5.47 | 3.89 | 6 | 28 | 41 | 13 | 100 |
| 52 | 5.45 | 3.88 | 6 | 28 | 40 | 13 | 100 |
| 53 | 5.43 | 3.87 | 6 | 28 | 40 | 13 | 100 |
| 54 | 5.41 | 3.85 | 6 | 27 | 39 | 13 | 100 |

| Age | Condition 1 | | Condition 2 | | | Condition 3 | |
|---|---|---|---|---|---|---|---|
| | Putamen | Caudate | TMS | SDMT[a] | SDMT[b] | TFC | IS |
| 55 | 5.39 | 3.84 | 6 | 27 | 39 | 13 | 100 |
| 56 | 5.38 | 3.83 | 6 | 26 | 38 | 13 | 100 |
| 57 | 5.36 | 3.81 | 6 | 26 | 38 | 13 | 100 |
| 58 | 5.34 | 3.80 | 6 | 25 | 37 | 13 | 100 |
| 59 | 5.32 | 3.79 | 7 | 25 | 37 | 12 | 100 |
| 60 | 5.31 | 3.77 | 7 | 24 | 36 | 12 | 99 |
| 61 | 5.29 | 3.76 | 8 | 24 | 36 | 12 | 99 |
| 62 | 5.27 | 3.75 | 8 | 23 | 35 | 12 | 99 |
| 63 | 5.25 | 3.73 | 8 | 22 | 35 | 12 | 99 |
| 64 | 5.24 | 3.72 | 8 | 22 | 34 | 12 | 98 |
| 65 | 5.22 | 3.71 | 9 | 21 | 33 | 12 | 98 |
| 66 | 5.20 | 3.70 | 9 | 21 | 33 | 12 | 97 |
| 67 | 5.18 | 3.68 | 9 | 20 | 32 | 12 | 97 |
| 68 | 5.17 | 3.67 | 9 | 19 | 31 | 12 | 97 |
| 69 | 5.15 | 3.66 | 10 | 18 | 31 | 12 | 96 |
| 70 | 5.13 | 3.64 | 10 | 18 | 30 | 11 | 96 |
| 71 | 5.11 | 3.63 | 11 | 17 | 29 | 11 | 95 |
| 72 | 5.09 | 3.62 | 11 | 16 | 29 | 11 | 94 |
| 73 | 5.08 | 3.60 | 11 | 16 | 28 | 11 | 94 |
| 74 | 5.06 | 3.59 | 12 | 15 | 27 | 11 | 93 |
| 75 | 5.04 | 3.58 | 12 | 14 | 26 | 11 | 92 |
| 76 | 5.02 | 3.56 | 13 | 13 | 25 | 11 | 92 |
| 77 | 5.01 | 3.55 | 13 | 12 | 25 | 10 | 91 |
| 78 | 4.99 | 3.54 | 14 | 12 | 24 | 10 | 90 |
| 79 | 4.97 | 3.53 | 14 | 11 | 23 | 10 | 90 |
| 80 | 4.95 | 3.51 | 14 | 10 | 22 | 10 | 89 |
| 81 | 4.94 | 3.50 | 15 | 9 | 21 | 10 | 88 |
| 82 | 4.92 | 3.49 | 15 | 8 | 21 | 10 | 87 |
| 83 | 4.90 | 3.47 | 16 | 8 | 20 | 10 | 87 |
| 84 | 4.88 | 3.46 | 16 | 7 | 19 | 9 | 86 |
| 85 | 4.87 | 3.45 | 17 | 6 | 18 | 9 | 85 |
| 86 | 4.85 | 3.43 | 17 | 5 | 17 | 9 | 84 |
| 87 | 4.83 | 3.42 | 18 | 4 | 16 | 9 | 83 |
| 88 | 4.81 | 3.41 | 18 | 3 | 16 | 9 | 83 |
| 89 | 4.80 | 3.39 | 19 | 2 | 15 | 9 | 82 |
| 90 | 4.78 | 3.38 | 20 | 2 | 14 | 8 | 81 |

[a] Less than or equal to high school education
[b] Greater than high school education

### G. Stage Characteristics

After scoring conditions and Stages for case visits, we evaluated the extent to which the condition indicators ($c_{hij}$) showed a monotone pattern at a visit. We also examined the extent to which the Stages showed ordered progression over visits for participants with repeated measures, focusing on the frequency distribution of the patterns.

#### i. Individual Visits

Table S9 shows the condition indicators for each visit along with the Stage assignment. The proportions in the table are computed by pooling the results among the imputed data sets (with 95% CIs). The upper portion shows the monotone patterns and the lower portion shows the non-monotone. The cumulative proportions indicate that 87% of the patterns are monotone, showing consistency with the HD-ISS.

***Table S9. Condition patterns with Stage assignment for individual visits.*** *Proportions and 95% CIs are based on multiple imputation. Upper half shows the monotone patterns consistent with the Staging System and the lower half shows the non-monotone.*

| $c_1$ | $c_2$ | $c_3$ | Assigned Stage | Proportion | Cumulative |
|---|---|---|---|---|---|
| 0 | 0 | 0 | 0 | 0.218 (0.203, 0.233) | 0.218 |
| 1 | 0 | 0 | 1 | 0.243 (0.227, 0.259) | 0.462 |
| 1 | 1 | 0 | 2 | 0.226 (0.210, 0.242) | 0.688 |
| 1 | 1 | 1 | 3 | 0.186 (0.170, 0.201) | 0.873 |
| 0 | 1 | 0 | 1 | 0.065 (0.056, 0.075) | 0.939 |
| 0 | 0 | 1 | 1 | 0.017 (0.012, 0.022) | 0.956 |
| 0 | 1 | 1 | 2 | 0.019 (0.013, 0.024) | 0.975 |
| 1 | 0 | 1 | 2 | 0.025 (0.019, 0.032) | 1.000 |

#### ii. Longitudinal Progression

To assess the progression of Stages for each participant, the ordinal correlation coefficient Kendall's tau-a was computed[35] for participants with at least two visits. Stage order was correlated with time order by comparing every pair of Stages with the associated pair of visit ages for a participant. In this context, tau-a is the proportion of monotonic-increasing Stage orderings (i.e., low to high) minus the proportion of incorrect Stage orderings (i.e., high to low). When there is no overall change in Stage, tau-a = 0; when there is more decrease than increase, tau-a < 0 (with lower limit -1); and when there is more increase than decrease, tau-a > 0 (with 1 being perfect ordering). Trends among all the cases were examined based on the frequency distribution of the individual tau-a correlations. To facilitate interpretation, we grouped the tau-a distribution into the categories of tau-a $< 0$, tau-a $= 0$, and tau-a $> 0$.

Table S10 shows the proportions for grouped tau-a values of individuals with different numbers of repeated visits (the proportions are the pooled results among the imputed data sets). The table indicates that the proportion of tau-a values consistent with correct Stage progression (i.e., tau-a $\geq 0$) varies from 86% to 89%. As the number of minimum visits increases, the proportion of tau-a > 0 also increases. This shows that individuals do tend to progress through increasing Stages given a sufficient time window.

***Table S10. Kendall's tau-a results as a function of number of visits.*** *Proportions and 95% CIs are pooled among multiple imputations.*

| Total $N$ | Visits | Tau-a | Proportion (95% CI) |
|---|---|---|---|
| 274 | >1 | <0 | 0.113 (0.002, 0.225) |
| | | 0 | 0.624 (0.551, 0.697) |
| | | >0 | 0.263 (0.161, 0.364) |
| 241 | >2 | <0 | 0.141 (0.024, 0.258) |
| | | 0 | 0.495 (0.406, 0.585) |
| | | >0 | 0.363 (0.263, 0.464) |
| 174 | >3 | <0 | 0.110 (0.000, 0.250) |
| | | 0 | 0.408 (0.293, 0.522) |
| | | >0 | 0.482 (0.375, 0.589) |
| 59 | >4 | <0 | 0.143 (0.000, 0.379) |
| | | 0 | 0.319 (0.108, 0.529) |
| | | >0 | 0.538 (0.365, 0.712) |

### H. Stage Probabilities

The final step focused on the estimation of the cumulative probability of transitioning into a Stage and the marginal probability of being in a Stage as a function of age and CAG expansion (for cases only). An ordinal regression approach was used that extends the continuation ratio (CR) model[36]fi to the longitudinal setting by including a random intercept[37].

#### i. Continuation Ratio Regression for Longitudinal Data

The CR model is useful for an ordinal variable in which increasing integer values represent transitions that occur in order from lower to higher disease states.[38,39] Such an ordering was hypothesized to be the case for the HD-ISS variable, $s_{ij}$.

In the longitudinal context, we are interested in the conditional probability of the $i^{th}$ participant being in Stage $h$ ($h = 1,2,3$) at time $j$ given they are already in Stage $h$ or a lower Stage. This is expressed as the CR,

$$\eta_{ij} = Pr(s_{ij} = h | s_{ij} \leq h) = \frac{Pr(s_{ij} = h)}{Pr(s_{ij} \leq h)}.$$

The response in the longitudinal ordinal regression model is the log of the CR odds, which is the log of the ratio of the probability of being in a particular Stage relative to the probability of being in a lower Stage,

$$log\left\{\frac{\eta_{ij}}{1-\eta_{ij}}\right\} = log\left\{\frac{Pr(s_{ij} = h)}{Pr(s_{ij} < h)}\right\}.$$

There are several options for the linear predictor of the outcome, and we considered five nested models. The most complex model, labeled as Model 5, provides maximal flexibility in the Stage curves. The intention is that if the data were incompatible with the HD-ISS, then Model 5 provided enough flexibility to reveal such incompatibilities. Model 5 is written as,

$$\begin{aligned} log\left\{\frac{\Pr(s_{ij}=h)}{\Pr(s_{ij}<h)}\right\} &= a_i + (\alpha_0 + \alpha_1\, I\{h=2\} + \alpha_2\, I\{h=1\}) \\ &+ age'_{ij}(\beta_0 + \beta_1\, I\{h=2\} + \beta_2\, I\{h=1\}) \\ &+ cag'_i(\gamma_0 + \gamma_1\, I\{h=2\} + \gamma_2\, I\{h=1\}) \\ &+ age'_{ij}cag'_i(\delta_0 + \delta_1\, I\{h=2\} + \delta_2\, I\{h=1\}), \end{aligned} \qquad \text{(A3)}$$

where $I\{\cdot\}$ is the indicator function (1 if the condition is true and 0 otherwise); $a_i$ is a random intercept; $\alpha_0$ is the fixed intercept for Stage 3, $\alpha_0 + \alpha_1$ is for Stage 2, and $\alpha_0 + \alpha_2$ is for Stage 1; $\beta_k$, $\gamma_k$, and $\delta_k$ are also fixed effects coefficients. To facilitate estimation, we set $age'_{ij} = age_{ij} - 18, cag'_i = cag_i - 40$.

The interactions involving the indicator variables allowed the Stage curves to vary flexibly in terms of location and slope by CAG. For the random intercept it is assumed that $a_i \sim \mathcal{N}(0, \sigma^2_{a_i})$, which means that Model 5 has 13 parameters.

The remaining models impose constraints on the Stage curves as a function of CAG. Model 4 sets $\delta_1 = \delta_2 = 0$ (11 parameters), so that the age by CAG interaction does not vary by Stage. Model 3 additionally sets $\beta_1 = \beta_2 = 0$ (9 parameters), and Model 2 additionally sets $\gamma_1 = \gamma_2 = 0$ (7 parameters). Finally, Model 1 additionally sets $\delta_0 = 0$ omitting the age by CAG interaction (6 parameters).

With the normality assumption for the random intercepts, ML methods were used for parameter estimation, and the Akaike Information Criterion (AIC)[40] was used for model evaluation. It can be shown that by re-coding the original data, software for general logistic regression analysis can be used for CR models,[41,42] which includes longitudinal models. We used the helper functions from the R package GLMMadaptive[19] for the re-coding and used the mixed_model() function to estimate the parameters with adaptive quadrature using a quasi-Newton algorithm.

The coefficients of Equation A3 are individual-specific. The goal of inference was to estimate Stage probabilities for a cohort of cases with the same CAG length, which required the population-averaged coefficients. Population-averaged coefficients were obtained by integrating with respect to the random intercepts. An approximation to the integration is possible based on quadrature methods[18], and the approximation was applied with the GLMMadaptive package. Thus, population averaged coefficients were used to depict the change in probabilities over time. Population averaged coefficients are denoted with an asterisk, e.g., $\alpha^*_0$.

Population-averaged parameter estimates of the five longitudinal ordinal regression models are listed in Table S11. MI Boot 95% CIs are shown in parentheses and the parameter estimates are listed in the logit metric (age and cag are each scaled, see above). Also included is the estimated intercept variance and the AIC, both based on the conditional random effects model.

The AIC shows a large decrease of 56 points going from Model 1 to Model 2, reflecting the importance of the age by CAG interaction. For Models 2-5, there is successive improvement in fit, but the improvement tends to diminish with model complexity ($\text{AIC}_2 - \text{AIC}_3 = 10; \text{AIC}_3 - \text{AIC}_4 = 13, \text{AIC}_4 - \text{AIC}_5 = 4$). Models 3-5 have several CIs that cover 0, whereas Model 1 and 2 have no CIs that cover 0.

Additional model comparison is provided in Figure S3 showing the predicted cumulative probability of transitioning into a Stage ($h \geq 1$) for cases with CAG = 40, 44, 48 (ribbons are the 95% MI Boot CIs). The panels indicate that the estimated models have similar predicted curves. The models with more parameters have greater CI widths because correlated parameters increase SEs. Notably, Model 5 has very similar predicted curves as Model 2 despite having 6 additional parameters. Given the relatively small AIC difference among models 2-5, the similarity of the predicted curves, and the CI coverage in Table S11, Model 2 was used to compute the marginal probabilities. In terms of the population-average coefficients, Model 2 is $log\left\{\frac{Pr(s_{ij}=h)}{Pr(s_{ij}<h)}\right\} = (\alpha_0^* + \alpha_1^* I\{h = 2\} + \alpha_2^* I\{h = 1\}) + \beta_0^* age'_{ij} + \gamma_0^* cag'_i + \delta_0^* age'_{ij} cag'_i$.

***Table S11. Population-averaged coefficients of the longitudinal continuation ratio models with 95% MI Boot CIs.*** *The variance of the intercepts and the AIC are based on the conditional random effects model.*

| Par | Model 1 | Model 2 | Model 3 | Model 4 | Model 5 |
|---|---|---|---|---|---|
| $a^*{}_0$ | -7.568<br>(-8.220, -6.917) | -6.624<br>(-7.298, -5.950) | -6.331<br>(-7.020, -5.641) | -7.797<br>(-9.008, -6.586) | -7.450<br>(-8.715, -6.185) |
| $a^*{}_1$ | 1.633<br>(1.476, 1.790) | 1.684<br>(1.519, 1.850) | 1.437<br>(1.181, 1.694) | 2.974<br>(1.836, 4.113) | 2.502<br>(1.291, 3.714) |
| $a^*{}_2$ | 3.570<br>(3.346, 3.793) | 3.631<br>(3.404, 3.859) | 3.379<br>(3.036, 3.722) | 5.521<br>(4.298, 6.743) | 5.179<br>(3.877, 6.481) |
| $b^*{}_0$ | 0.151<br>(0.135, 0.167) | 0.113<br>(0.094, 0.131) | 0.109<br>(0.090, 0.127) | 0.148<br>(0.117, 0.178) | 0.136<br>(0.103, 0.170) |
| $g^*{}_0$ | 0.643<br>(0.566, 0.719) | 0.249<br>(0.112, 0.386) | 0.143<br>(-0.022, 0.307) | 0.296<br>(0.102, 0.491) | 0.144<br>(-0.134, 0.422) |
| $d^*{}_0$ | | 0.018<br>(0.012, 0.024) | 0.020<br>(0.014, 0.027) | 0.019<br>(0.013, 0.025) | 0.024<br>(0.015, 0.034) |
| $g^*{}_1$ | | | 0.086<br>(0.013, 0.159) | -0.049<br>(-0.173, 0.074) | 0.168<br>(-0.092, 0.427) |
| $g^*{}_2$ | | | 0.092<br>(-0.017, 0.200) | -0.114<br>(-0.273, 0.046) | 0.036<br>(-0.271, 0.343) |
| $b^*{}_1$ | | | | -0.041<br>(-0.069, -0.013) | -0.025<br>(-0.057, 0.007) |
| $b^*{}_2$ | | | | -0.060<br>(-0.090, -0.030) | -0.049<br>(-0.085, -0.014) |
| $d^*{}_1$ | | | | | -0.009<br>(-0.018, 0.000) |
| $d^*{}_2$ | | | | | -0.006<br>(-0.018, 0.007) |

| Par | Model 1 | Model 2 | Model 3 | Model 4 | Model 5 |
|---|---|---|---|---|---|
| $s^2_a$ | 7.354 | 7.096 | 7.222 | 7.261 | 7.264 |
| | (5.891, 8.817) | (5.689, 8.504) | (5.746, 8.698) | (5.751, 8.771) | (5.740, 8.788) |
| AI | 6255.0 | 6199.1 | 6188.9 | 6176.1 | 6171.9 |
| C | (5989.6, 6520.4) | (5932.6, 6465.5) | (5921.7, 6456.1) | (5908.8, 6443.4) | (5904.5, 6439.2) |

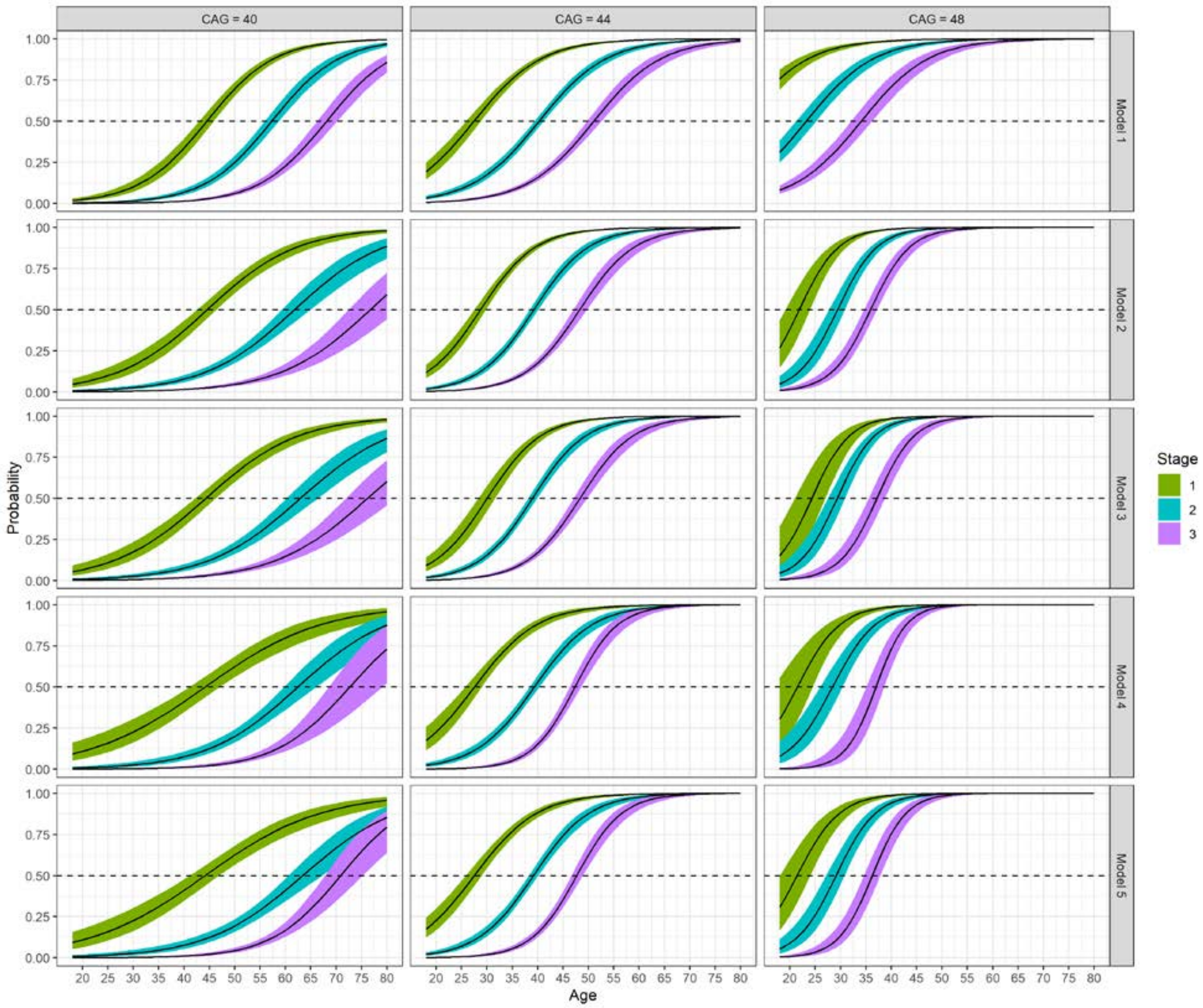


***Figure S3. Predicted cumulative probability curves (95% MI Boot CIs) for five longitudinal models paneled by CAG.*** *The curves represent the probability of transitioning into a Stage from a lower Stage over time.*

Figure 3 in the main text shows the estimated probabilities and Table 2 lists representative values. Using population-averaged coefficients, we computed the predicted probability of transitioning into the $h^{th}$ Stage ($h \geq 1$) over time for a cohort. Formulas for computing the marginal probabilities from Model 2 are the following. For Model 2 based on Equation 2, let $\phi_{ij} = \beta_0 age_{ij} + \gamma_0 cag_i + \delta_0 age_{ij} cag_i$. Then the marginal probabilities are,

$$Pr(s_{ij} = 3) = \frac{exp(\alpha_0 + \phi_{ij} + a_i)}{1 + exp(\alpha_0 + \phi_{ij} + a_i)},$$

$$Pr(s_{ij} = 2) = \frac{exp(\alpha_0 + \alpha_1 + \phi_{ij} + a_i)}{1 + exp(\alpha_0 + \alpha_1 + \phi_{ij} + a_i)} \cdot \frac{1}{1 + exp(\alpha_0 + \phi_{ij} + a_i)},$$

$$Pr(s_{ij} = 1) = \frac{exp(\alpha_0 + \alpha_2 + \phi_{ij} + a_i)}{1 + exp(\alpha_0 + \alpha_2 + \phi_{ij} + a_i)} \cdot \frac{1}{1 + exp(\alpha_0 + \alpha_1 + \phi_{ij} + a_i)} \cdot \frac{1}{1 + exp(\alpha_0 + \phi_{ij} + a_i)},$$

$$Pr(s_{ij} = 0) = 1 - \sum_{h>1} Pr(s_{ij} = h).$$

Similar relations hold when considering the population-average coefficients (omitting the random intercept).

### I. Mapping Clinical Variables to Stage Probabilities

Clinical variables were mapped to the stage probabilities as shown in Figure 5 in the main text. Linear mixed models with cubic splines for age were used to fit smooth curves for the clinical variables (each model had a random intercept and random slope). Predicted values were computed based on the estimated fixed effects. The scaled curves in Figure 5A (0-1 metric) were computed by calibrating to curves computed for controls. The curves in Figure 5A are the proportion differences from the control at each age.

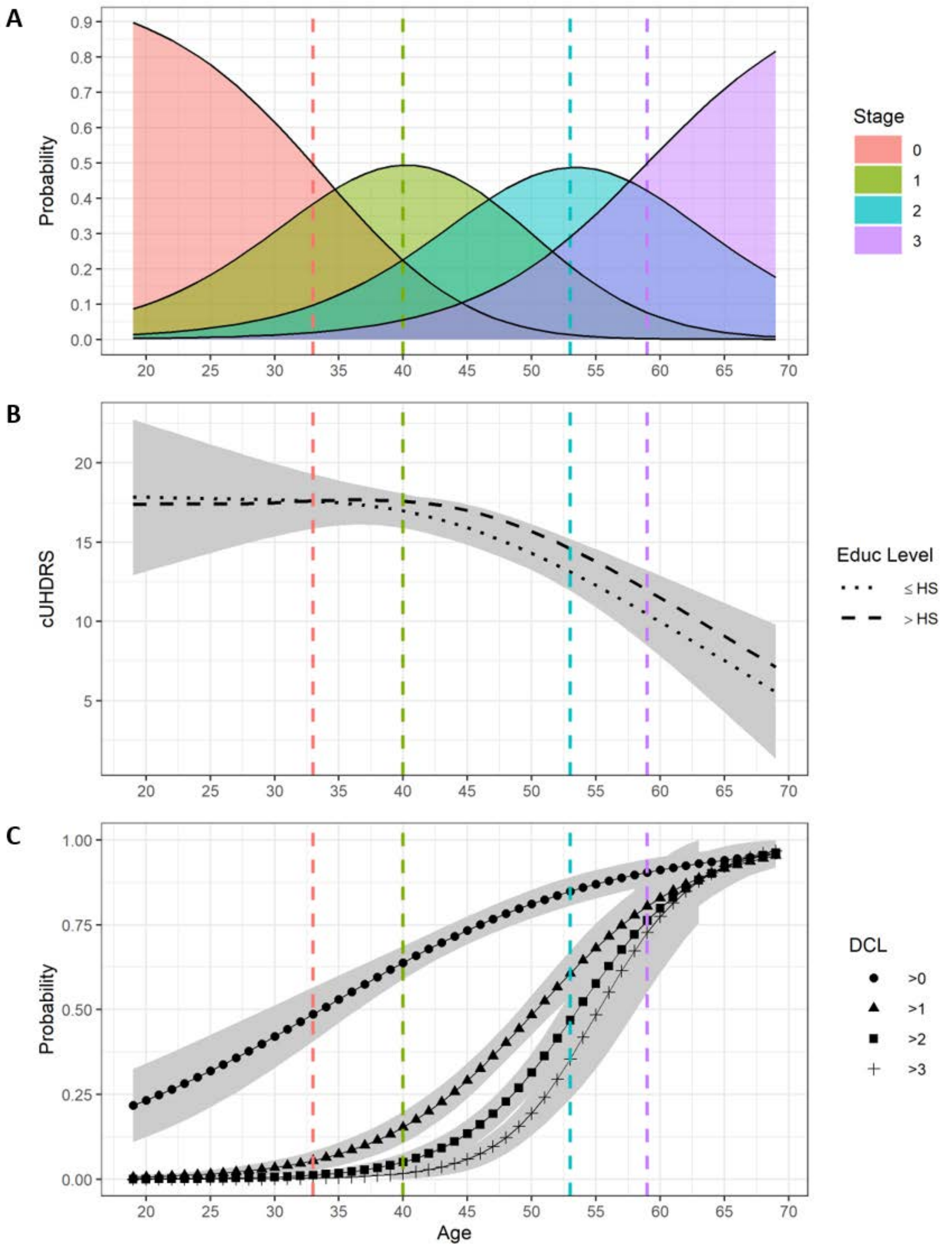


***Figure S4. Correspondence of the cUHDRS and DCL to the HD-ISS for CAG = 42.*** *Panel A shows the Stage probabilities as a function of age. Panel B shows predicted mean cUHDRS, with two education levels for SDMT (ed ≤ HS, ed > HS). Panel C shows the predicted mean DCL. In Panels B-C a vertical dashed line (stage colors as in Panel A) denotes the age at which a Stage probability = 0.50.*

### 5. vMRI Technical Details

We converted the imaging data from DICOM to Nifti format using the dcm2niix tool. Subsequently, we performed full brain gray matter segmentation using the recon-all pipeline from Freesurfer v6 (build freesurfer-Linux-centos6_x86_64-stable-pub-v6.0.1-f53a55a). To reduce the noise in the segmentation data, we followed the longitudinal pipeline which consists in a first round of cross-sectional segmentation, the creation of a subject-specific template (SST) from all the timepoints, and the re-segmentation of each longitudinal timepoint using the SST template prior (http://dx.doi.org/10.1016/j.neuroimage.2012.02.084). The volumetric scores were obtained from the Desikan-Killiany-Tourville atlas at each timepoint (file aparc.DKTatlas+aseg.mgz). All Freesurfer processing was conducted in a cloud computing environment which included multiple servers.

To improve the quality of the research, we performed visual quality control (QC) inspections of the Freesurfer segmentations. During this step, we checked the overall plausibility of the segmentation and focused more specifically on the accuracy of the caudate and putamen segmentations. We noticed that occasional failures in the first round of cross-sectional segmentation would propagate the error in the second round of longitudinal segmentation. Since failures were more easily identifiable in the first round of segmentation, we performed the QC only on the cross-sectional segmentations. During the QC, each segmentation was assigned a numeric value in a 5-point rating scale; a binary cut-off was used subsequently to exclude scores derived from failed segmentations (1-2 = fail, 3-5 = pass). The typical failure rate of the segmentation was ~5% of the timepoints, and the most typical reason of failure was an under segmentation of the putamen. A systematic error was observed throughout all the data – partial inclusion of the nearby claustrum in the putamen label, but this issue was deemed to be minor and unavoidable.

### 6. Supplementary Figure S5

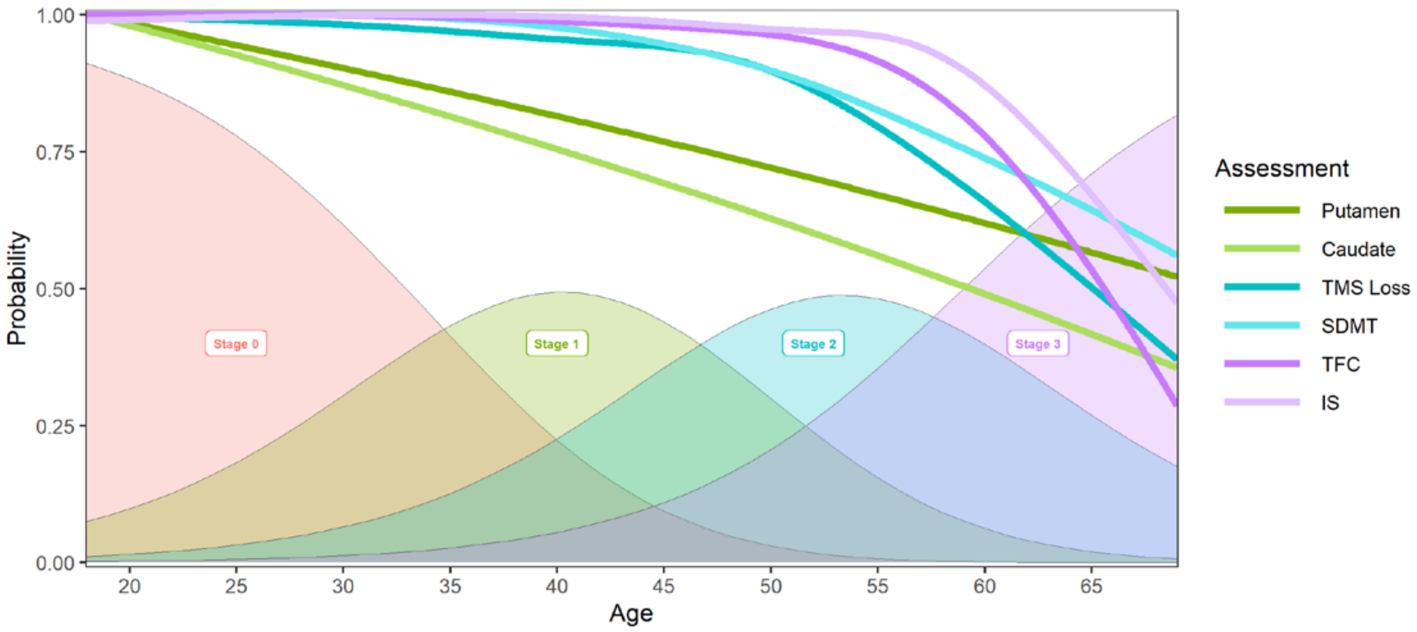


***Figure S5. Overview of the HD-ISS for CAG = 42.*** *Stage probabilities (shaded areas) and unit-scaled mean predicted curves as a function of age. The predicted means were scaled relative to the control curves and represent the proportion difference from the controls. The TMS was reverse scored prior to the scaling and represents loss from normal functioning.*

### 7. References


1 Wijeratne, P. A. *et al.* An image-based model of brain volume biomarker changes in Huntington's disease. *Ann Clin Transl Neurol* **5**, 570-582, doi:10.1002/acn3.558 (2018).

2 Wijeratne, P. A. *et al.* Robust Markers and Sample Sizes for Multicenter Trials of Huntington Disease. *Ann Neurol* **87**, 751-762, doi:10.1002/ana.25709 (2020).

3 Sun, Z. *et al.* A Data-Driven Method for Generating Robust Symptom Onset Indicators in Huntington's Disease Registry Data. *AMIA Annu Symp Proc* **2017**, 1635-1644 (2017).

4 Ghosh, S. *et al.* An Exploration of Latent Structure in Observational Huntington's Disease Studies. *AMIA Jt Summits Transl Sci Proc* **2017**, 92-102 (2017).

5 Tabrizi, S. J. *et al.* Biological and clinical manifestations of Huntington's disease in the longitudinal TRACK-HD study: cross-sectional analysis of baseline data. *Lancet Neurol* **8**, 791-801, doi:10.1016/S1474-4422(09)70170-X (2009).

6 Paulsen, J. S. *et al.* Prediction of manifest Huntington's disease with clinical and imaging measures: a prospective observational study. *Lancet Neurol* **13**, 1193-1201, doi:10.1016/S1474-4422(14)70238-8 (2014).

7 Tabrizi, S. J. *et al.* Predictors of phenotypic progression and disease onset in premanifest and early-stage Huntington's disease in the TRACK-HD study: analysis of 36-month observational data. *Lancet Neurol* **12**, 637-649, doi:10.1016/S1474-4422(13)70088-7 (2013).

8 Long, J. D., Paulsen, J. S., Investigators, P.-H. & Coordinators of the Huntington Study, G. Multivariate prediction of motor diagnosis in Huntington's disease: 12 years of PREDICT-HD. *Mov Disord* **30**, 1664-1672, doi:10.1002/mds.26364 (2015).

9 Li, F. *et al.* Predicting the Risk of Huntington's Disease with Multiple Longitudinal Biomarkers. *J Huntingtons Dis* **8**, 323-332, doi:10.3233/JHD-190345 (2019).

10 Paulsen, J. S. *et al.* Challenges assessing clinical endpoints in early Huntington disease. *Mov Disord* **25**, 2595-2603, doi:10.1002/mds.23337 (2010).

11 Beglinger, L. J. *et al.* Clinical predictors of driving status in Huntington's disease. *Mov Disord* **27**, 1146-1152, doi:10.1002/mds.25101 (2012).

12 Griffiths, A. J. F., J. H. Miller, D. T. Suzuki, R. C. Lewontin, and W. M. Gelbart. . *An Introduction to Genetic Analysis*. 7th Edition edn, (W. H. Freeman, 2000).

13 United Nations. *World Population Prospects 2019*, <https://population.un.org/wpp/> (2019).

14 Landwehrmeyer, G. B. *et al.* Data Analytics from Enroll-HD, a Global Clinical Research Platform for Huntington's Disease. *Mov Disord Clin Pract* **4**, 212-224, doi:10.1002/mdc3.12388 (2017).

15 Paulsen, J. S. *et al.* Preparing for preventive clinical trials: the Predict-HD study. *Arch Neurol* **63**, 883-890, doi:10.1001/archneur.63.6.883 (2006).

16 Paulsen, J. S. *et al.* Detection of Huntington's disease decades before diagnosis: the Predict-HD study. *J Neurol Neurosurg Psychiatry* **79**, 874-880, doi:10.1136/jnnp.2007.128728 (2008).

17 Paulsen, J. S. *et al.* Clinical and Biomarker Changes in Premanifest Huntington Disease Show Trial Feasibility: A Decade of the PREDICT-HD Study. *Front Aging Neurosci* **6**, 78, doi:10.3389/fnagi.2014.00078 (2014).

18 Hedeker, D., du Toit, S. H. C., Demirtas, H. & Gibbons, R. D. A note on marginalization of regression parameters from mixed models of binary outcomes. *Biometrics* **74**, 354-361, doi:10.1111/biom.12707 (2018).

19 Rizopoulos, D. *GLMMadaptive: Generalized Linear Mixed Models Using Adaptive Gaussian Quadrature*, <https://CRAN.R-project.org/package=GLMMadaptive> (2020).
20 Georgiou-Karistianis, N. *et al.* Automated differentiation of pre-diagnosis Huntington's disease from healthy control individuals based on quadratic discriminant analysis of the basal ganglia: the IMAGE-HD study. *Neurobiol Dis* **51**, 82-92, doi:10.1016/j.nbd.2012.10.001 (2013).
21 Kloppel, S. *et al.* Compensation in Preclinical Huntington's Disease: Evidence From the Track-On HD Study. *EBioMedicine* **2**, 1420-1429, doi:10.1016/j.ebiom.2015.08.002 (2015).
22 Mills, J. A., Long, J. D., Mohan, A., Ware, J. J. & Sampaio, C. Cognitive and Motor Norms for Huntington's Disease. *Arch Clin Neuropsychol* **35**, 671-682, doi:10.1093/arclin/acaa026 (2020).
23 Koenker, R. Quantile Regression for Longitudinal Data. *Journal of Multivariate Analysis* **91**, 74-89 (2004).
24 Koenker, R., and S. H. Bache. *Rqpd: Regression Quantiles for Panel Data.*, <https://R-Forge.R-project.org/projects/rqpd/> (2014).
25 Corr, T. E., Schaefer, E. W. & Paul, I. M. Growth during the first year in infants affected by neonatal abstinence syndrome. *BMC Pediatr* **18**, 343, doi:10.1186/s12887-018-1327-0 (2018).
26 Koscik, R. L. *et al.* Longitudinal Standards for Mid-life Cognitive Performance: Identifying Abnormal Within-Person Changes in the Wisconsin Registry for Alzheimer's Prevention. *J Int Neuropsychol Soc* **25**, 1-14, doi:10.1017/S1355617718000929 (2019).
27 Ounpraseuth, S. T., Magann, E. F., Spencer, H. J., Rabie, N. Z. & Sandlin, A. T. Normal amniotic fluid volume across gestation: Comparison of statistical approaches in 1190 normal amniotic fluid volumes. *J Obstet Gynaecol Res* **43**, 1122-1131, doi:10.1111/jog.13332 (2017).
28 Wei, Y., Pere, A., Koenker, R. & He, X. Quantile regression methods for reference growth charts. *Stat Med* **25**, 1369-1382, doi:10.1002/sim.2271 (2006).
29 Rubin, D. B. *Multiple Imputation for Nonresponse in Surveys*. (John Wiley & Sons., 1987).
30 Little, R. J. A. Missing-Data Adjustments in Large Surveys. *Journal of Business & Economic Statistics* **6**, 287–296 (1988).
31 Breiman, L. Random Forests. *Machine Learning* **45**, 5-32, doi:https://doi.org/10.1023/A:1010933404324 (2001).
32 Morris, T. P., White, I. R. & Royston, P. Tuning multiple imputation by predictive mean matching and local residual draws. *BMC Med Res Methodol* **14**, 75, doi:10.1186/1471-2288-14-75 (2014).
33 Schomaker, M. & Heumann, C. Bootstrap inference when using multiple imputation. *Stat Med* **37**, 2252-2266, doi:10.1002/sim.7654 (2018).
34 Mayer, M. *MissRanger: Fast Imputation of Missing Values*, <https://CRAN.R-project.org/package=missRanger> (2019).
35 Kendall, M. G., and Gibbons, J. D. . *Rank Correlation Methods*. 5th edn, (Oxford, 1990).
36 Fienberg, S. E. *The Analysis of Cross-Classified Categorical Data*. . (MIT Press, 1977).
37 Dos Santos, D. M. & Berridge, D. M. A continuation ratio random effects model for repeated ordinal responses. *Stat Med* **19**, 3377-3388, doi:10.1002/1097-0258(20001230)19:24<3377::aid-sim526>3.0.co;2-e (2000).

38 Harrell, F. E. *Regression Modeling Strategies.*, (Springer, 2015).
39 Meisner, A., Parikh, C. R. & Kerr, K. F. Using ordinal outcomes to construct and select biomarker combinations for single-level prediction. *Diagn Progn Res* **2**, 8, doi:10.1186/s41512-018-0028-3 (2018).
40 Akaike, H. in *Second International Symposium on Information Theory* (ed B. N. Petrov and F. Csaki) 267–281 (Akademiai Kiado, 1973).
41 Berridge, D. M. & Whitehead, J. Analysis of failure time data with ordinal categories of response. *Stat Med* **10**, 1703-1710, doi:10.1002/sim.4780101108 (1991).
42 Armstrong, B. G. & Sloan, M. Ordinal regression models for epidemiologic data. *Am J Epidemiol* **129**, 191-204, doi:10.1093/oxfordjournals.aje.a115109 (1989).
